# Mapping disease regulatory circuits at cell-type resolution from single-cell multiomics data

**DOI:** 10.1101/2022.12.06.22282077

**Authors:** Xi Chen, Yuan Wang, Antonio Cappuccio, Wan-Sze Cheng, Frederique Ruf Zamojski, Venugopalan D. Nair, Clare M. Miller, Aliza B. Rubenstein, German Nudelman, Alicja Tadych, Chandra L. Theesfeld, Alexandria Vornholt, Mary-Catherine George, Felicia Ruffin, Michael Dagher, Daniel G. Chawla, Alessandra Soares-Schanoski, Rachel R. Spurbeck, Lishomwa C. Ndhlovu, Robert Sebra, Steven H. Kleinstein, Andrew G. Letizia, Irene Ramos, Vance G. Fowler, Christopher W. Woods, Elena Zaslavsky, Olga G. Troyanskaya, Stuart C. Sealfon

## Abstract

Resolving chromatin remodeling-linked gene expression changes at cell type resolution is important for understanding disease states. We describe MAGICAL, a hierarchical Bayesian approach that leverages paired scRNA-seq and scATAC-seq data from different conditions to map disease-associated transcription factors, chromatin sites, and genes as regulatory circuits. By simultaneously modeling signal variation across cells and conditions in both omics data types, MAGICAL achieved high accuracy on circuit inference. We applied MAGICAL to study *Staphylococcus aureus* sepsis from peripheral blood mononuclear single-cell data that we generated from infected subjects with bloodstream infection and from uninfected controls. MAGICAL identified sepsis-associated regulatory circuits predominantly in CD14 monocytes, known to be activated by bacterial sepsis. We addressed the challenging problem of distinguishing host regulatory circuit responses to methicillin-resistant-(MRSA) and methicillin-susceptible *Staphylococcus aureus* (MSSA) infections. While differential expression analysis failed to show predictive value, MAGICAL identified epigenetic circuit biomarkers that distinguished MRSA from MSSA.

## Introduction

Gene expression can be modulated through the interplay of proximal and distal regulatory domains brought together in 3D space^1^. Chromatin regulatory domains, transcription factors, and downstream target genes form regulatory circuits^2^. Within circuits, the binding of transcription factors to chromatin regions and the 3D looping between these regions and gene promoters represent the mechanisms governing how transcription factors transform regulatory signals into changes in RNA transcription^3, 4^. In disease, these circuits could be dysregulated in a cell type specific manner and may not be observed from bulk samples^5^. Therefore, identifying the impact of disease on regulatory circuits requires a framework for mapping regulatory domains with chromatin accessibility changes to altered gene expression in the context of cell-type resolution^6^. Single-cell RNA sequencing (scRNA-seq) and single-cell assay for transposase-accessible chromatin using sequencing (scATAC-seq) characterizing disease states have improved the identification of differential chromatin sites and/or differentially expressed genes within individual cell types^5, 7, 8^.

Yet, advances in single-cell assay technology have outpaced the development of methods to maximize the value of multiomics datasets for studying disease-associated regulation, especially for the regulatory interactions that are not directly measured by the omics data. Recent computational approaches^9–12^ to support the multiomics data analysis demonstrate the promise of this area but still lack the capacity to resolve regulation changes within individual cell types, which precludes elucidating regulatory circuits affected by the disease or showing different responses in varying disease states.

To address these, we developed MAGICAL (Multiome Accessibility Gene Integration Calling And Looping), a method that models coordinated chromatin accessibility and gene expression variation to identify circuits (both the units and their interactions) that differ between conditions. MAGICAL analyzes scRNA-seq and scATAC-seq data using a hierarchical Bayesian framework. To accurately detect differences in regulatory circuit activity between conditions, MAGICAL introduces hidden variables for explicitly modeling the transcriptomic and epigenetic signal variations between conditions and optimization against the noise in both scRNA-seq and scATAC-seq datasets. Because regulatory circuits are cell-type specific^13^, MAGICAL reconstructs them at cell-type resolution. Systematic benchmarking against multiple public datasets supported the accuracy of MAGICAL-identified regulatory circuits.

*Staphylococcus aureus* (*S. aureus*), a bacterium often resistant to common antibiotics, is a major cause of severe infection and mortality^14, 15^. Using single-cell multiomics data generated from peripheral blood mononuclear cell (PBMC) samples of *S. aureus* infected subjects and healthy controls, MAGICAL identified host response regulatory circuits that are modulated during *S. aureus* bloodstream infection, and circuits that discriminate the responses to methicillin-resistant (MRSA) and methicillin-susceptible *S. aureus* (MSSA). Genes in the host circuits accurately predicted *S. aureus* infection in multiple validation datasets. Moreover, in contrast to conventional differential analysis that failed to identify specific genes for robust antibiotic-sensitivity prediction, MAGICAL-identified circuit genes can differentiate MRSA from MSSA. Therefore, MAGICAL can be used for multiomics data-based gene signature development, providing a bioinformatic solution that can improve disease diagnosis.

## Results

### MAGICAL framework

MAGICAL identifies disease-associated regulatory circuits by comparing single-cell multiomics data (scRNA-seq and scATAC-seq) from disease and control samples (**Figure 1a**). The framework incorporates transcription factor (TF) motifs and chromatin topologically associated domain (TAD) boundaries as prior information to infer regulatory circuits comprising chromatin regulatory sites, modulatory TFs, and downstream target genes for each cell type. In brief, to build candidate disease-modulated circuits, differentially accessible sites (DAS) within each cell type are first associated with TFs by motif sequence matching and then linked to differentially expressed genes (DEG) in that cell type by genomic localization within the same TAD. Next, MAGICAL uses a Bayesian framework to iteratively model chromatin accessibility and gene expression variation across cells and samples in each cell type and to estimate the confidence of TF-peak and peak-gene linkages for each candidate circuit (**Figure 1b**).

**Figure 1:**
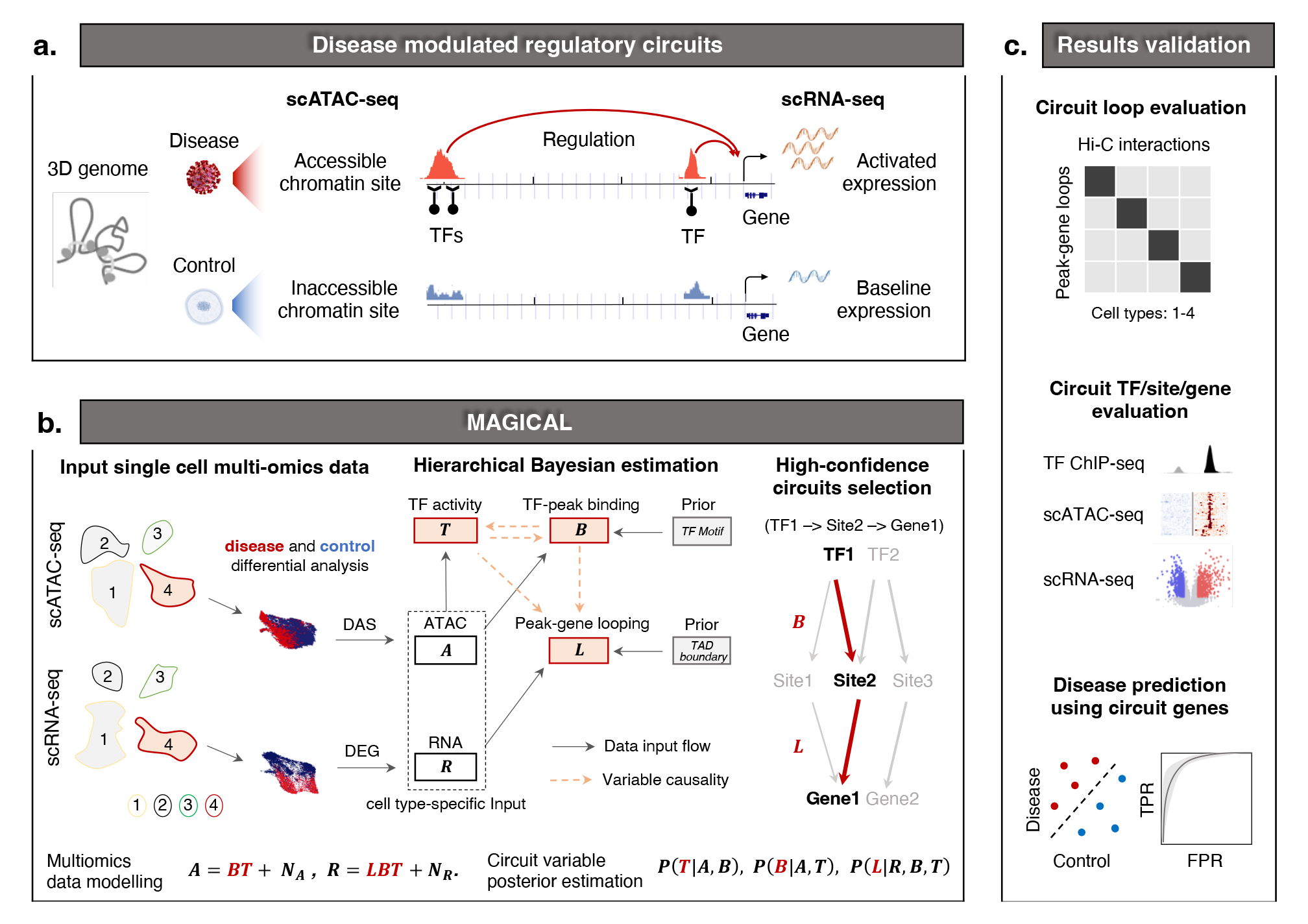
Overview of MAGICAL for mapping disease-associated regulatory circuits from scRNA-seq and scATAC-seq data. (**a**) In the 3D genome, the altered gene expression in cells between disease and control conditions can be attributed to the chromatin accessibility changes of proximal and distal chromatin sites regulated by TFs. (**b**) To identify disease-associated regulatory circuits in a selected cell type (including ATAC assay cells and RNA assay cells from samples being compared), MAGICAL selects DAS as candidate regions and DEG as candidate genes. Then, the filtered ATAC data and RNA data of DAS and DEG are used as input to a hierarchical Bayesian framework pre-embedded with the prior TF motifs and TAD boundaries. The chromatin activity ***A*** is modelled as a linear combination of TF-peak binding confidence ***B*** and the hidden TF activity ***T***, with contamination of data noise ***N_A_***. The gene expression ***R*** is modelled as a linear combination of ***B***, ***T***, and peak-gene looping confidence ***L***, with contamination of data noise ***N_R_***. MAGICAL estimates the posterior probabilities ***P***(***B***|***A***,***T***), ***P***(***T***|***A***,***B***) and ***P***(***L***|***R***,***B***,***T***) by iteratively sampling variables ***B***, ***T***, and ***L*** to optimize against the data noise ***N_A_*** and ***N_R_*** in both modalities. Finally, regulatory circuits with high posterior probabilities of ***B*** and ***L*** (e.g., a high confidence circuit with inferred interactions between TF1, Site2 and Gene1) are selected. (**c**) We evaluate the accuracy and cell type specificity of the inferred peak-gene looping interactions by checking their enrichment with cell-type matched chromatin interactions in Hi-C experiments. For the identified TFs, peaks, and genes in circuits, we checked the accuracy of each using independent ChIP-seq, scATAC-seq, and scRNA-seq data. Finally, as a demonstration of the utility of MAGICAL, we used the circuit target genes as features to predict disease states.

To accurately identify varying circuits between different conditions, MAGICAL explicitly models signal and noise in chromatin accessibility and gene expression data (Methods: MAGICAL). A TF-peak binding variable and a hidden TF activity variable are jointly estimated to fit to the chromatin accessibility variation across cells from the conditions being compared. These two variables are then used together with a peak-gene looping variable to fit the gene expression variation. Using Gibbs sampling, MAGICAL iteratively estimates variable values and optimizes the states of circuit TF-peak-gene linkages. Finally, high-confidence circuits fitting the signal variation in both data types are selected.

TF activity represents the regulatory capacity (protein level) of a particular TF protein^16, 17^, which is distinct from TF expression. For each TF, we assume its hidden TF activities following an identical distribution across cells in the same cell type and the same sample, regardless of if the cells are from the scATAC-seq assay or the scRNA-seq assay or both. MAGICAL iteratively learns the activity distribution for each TF and estimates the specific activities of all TFs in each cell (**Supplementary Figure 1**). This procedure eliminates the requirement of cell-level pairing of RNA-seq and ATAC-seq data. It makes MAGICAL a general tool that can analyze single-cell true multiome or sample-paired multiomics datasets.

We validated MAGICAL in multiple ways, demonstrating that it infers regulatory circuits accurately (**Figure 1c**). MAGICAL-inferred linkages between chromatin sites and genes were validated using experimental 3D chromatin interactions. The resulting circuit genes, peaks and their regulatory TFs were respectively evaluated in multiple independent studies. And finally, as one example of utility, we showed that the circuit genes can be used as features to classify disease states, providing a bioinformatics solution to challenging diagnostic problems.

### Comparative analysis of performance

MAGICAL is a scalable framework. It can infer regulatory circuits of TFs, chromatin regions, and genes with differential activities between contrast conditions or infer regulatory circuits with active chromatin regions and genes in a single condition. Because existing integrative methods^11, 12, 18^ can only be applied to single-condition data, to provide a comparative assessment of the performance of MAGICAL, we restricted MAGICAL to the single-condition data analysis possible with existing methods.

For peak-gene looping inference, we compared MAGICAL to the TRIPOD^11^ and FigR^18^ methods, using the same benchmark single-cell multiome datasets as used by the authors reporting these methods. In the comparison of MAGICAL with TRIPOD using a 10X multiome single-cell dataset, MAGICAL-inferred peak-gene loops showed significantly higher enrichment of experimentally observed chromatin interactions in blood cells in the 4DGenome database^19^ (*p-*value<0.0001, two-side Fisher’s exact test, **Supplementary Figure 2a**), the same validation data used by TRIPOD developers. MAGICAL also significantly outperformed FigR on the application to a GM12878 SHARE-seq dataset^10^. In that case, the peak-gene loops in MAGICAL-selected circuits had significantly higher enrichment of H3K27ac-centric chromatin interactions^20^ than did FigR (*p*-value<0.0001, two-side Fisher’s exact test, **Supplementary Figure 2b**).

Because the MAGICAL framework, unlike TRIPOD and FigR, used chromatin TAD boundaries as prior information, we evaluated whether the improvement in performance resulted solely from this additional information. To investigate this, we eliminated the use of TAD boundaries and modified MAGICAL for this test by assigning candidate linkages between peaks and genes within 500Kb (a naïve distance prior). As shown in **Supplementary Figures 2a** and **2b,** even without the TAD prior information, MAGICAL still outperformed the competing methods (*p-*values <0.001, two-side Fisher’s exact test). Overall, these results suggest that in addition to the benefit of priors, explicit modeling of signal and noise in both chromatin accessibility and gene expression data increased the accuracy of peak-gene looping identification.

**Figure 2.**
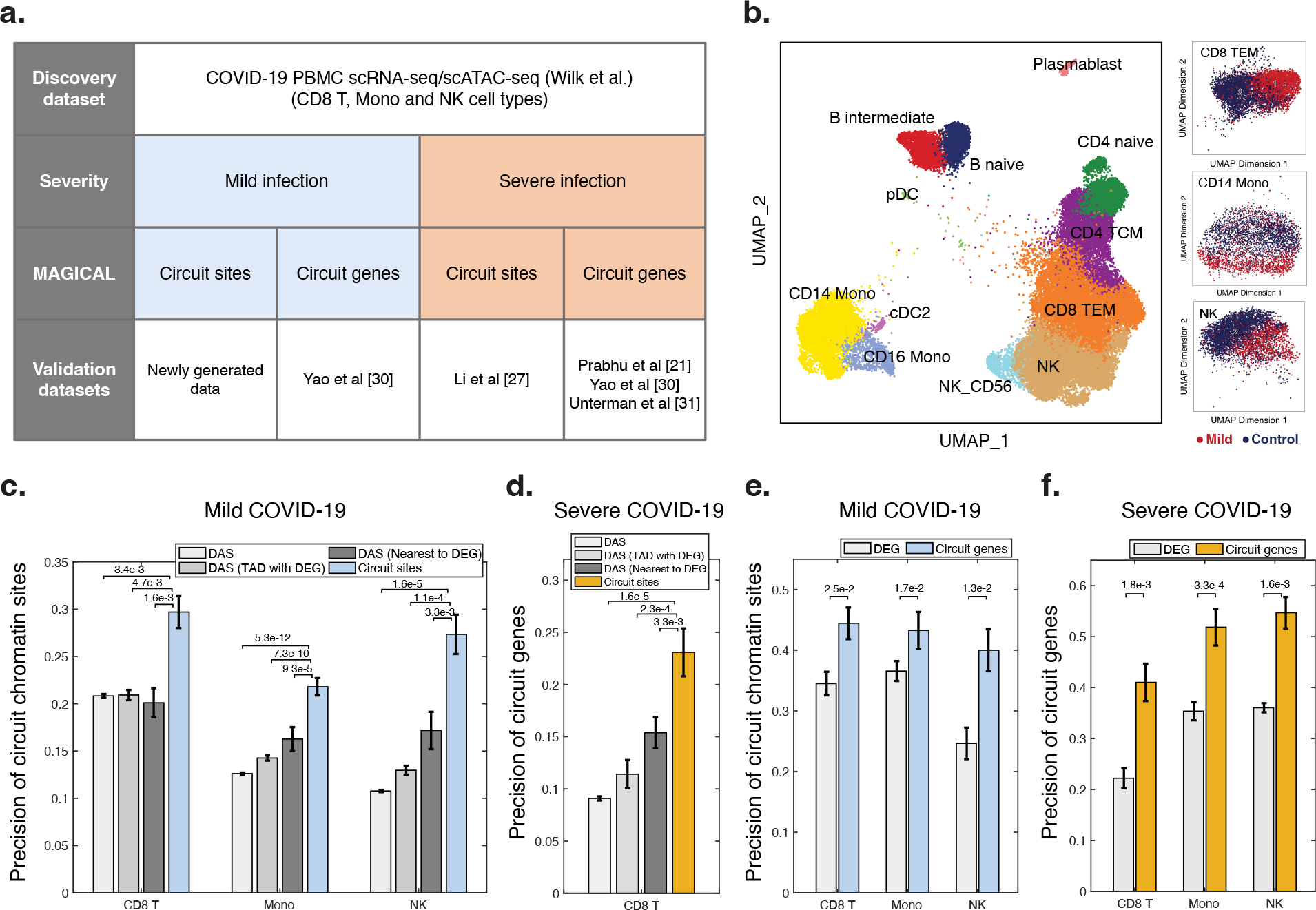
Validation of COVID-19-associated circuit chromatin sites and genes. (**a**) We applied MAGICAL to a COVID-19 PBMC single-cell multiomics dataset and identified circuits for the clinical mild and severe groups, respectively. We validated the circuit-associated chromatin sites and genes using newly generated and independent COVID-19 single-cell datasets. (**b**) UMAPs of a newly generated independent scATAC-seq dataset including 16K cells from 6 COVID-19 subjects and 9K cells from 3 controls showed chromatin accessibility changes in CD8 TEM, CD14 Mono, and NK cell types. (**c, d**) The precision of MAGICAL-selected circuit sites is significantly higher than the that of the original DAS, the nearest DAS to DEG or all DAS in the same TAD with DEG. (**e, f**) The precision of circuit genes are significantly higher than the that of DEG. (**c, e**) For mild COVID-19, MAGICAL identified 645 sites in CD8 TEM, 599 sites in CD14 Mono and 148 sites in NK, regulating 153 genes, 183 genes and 60 genes, respectively. (**d, f**) For severe COVID-19, MAGICAL identified 78 sites, 202 sites and 62 sites in the three cell types, regulating 25 genes, 81 genes, and 26 genes, respectively. (**c-f**) Precision is defined as the proportion of the identified circuit sites/genes to be differentially accessible and differentially expressed in the same cell type between infection and control conditions in independent datasets. Results are presented as bar plots where the height represent the precision and the error bar represent the 95% confidence interval. Significance evaluation is done using two-side Fisher’s exact test.

### MAGICAL analysis of COVID-19 single-cell multiomics data

To demonstrate the accuracy of the primary application of MAGICAL on contrast condition data to infer disease-modulated circuits, we applied MAGICAL to sample-paired peripheral blood mononuclear cell (PBMC) scRNA-seq and scATAC-seq data from SARS-CoV-2 infected individuals and healthy controls^5^. Because immune responses in COVID-19 patients differ according to disease severity^21, 22^, MAGICAL inferred the regulatory circuits for mild and severe clinical groups separately. The chromatin sites and genes in the identified circuits were validated using newly generated and publicly available independent COVID-19 single-cell datasets (**Figure 2a**). We primarily focused on three cell types that have been found to show widespread gene expression and chromatin accessibility changes in response to SARS-CoV-2 infection^23, 24^, including CD8 effector memory T (TEM) cells, CD14 monocytes (Mono), and natural killer (NK) cells. In total, MAGICAL identified 1,489 high confidence circuits (1,404 sites and 391 genes) in these cell types for mild and severe clinical groups (**Supplementary Table 1**; Methods: MAGICAL analysis of COVID-19 single-cell multiomics data).

To confirm the circuit chromatin sites selected by MAGICAL for mild COVID-19, we generated an independent PBMC scATAC-seq dataset from six SARS-CoV-2-infected subjects with mild symptoms and three uninfected (PCR-negative) controls (**Figure 2b**; **Supplementary Table 2**). About 25,000 quality cells were selected after quality-control (QC) analysis. These cells were integrated, clustered and annotated using ArchR^25^ (**Supplementary Figure 3; Supplementary Table 3**). Peaks were called from each cell type using MACS2^26^. In total, 284,909 peaks were identified (**Supplementary Table 4**). For the three selected cell types, differential analysis between COVID-19 and control returned 3,061 sites for CD8 TEM, 1,301 sites for CD14 Mono, and 1,778 sites for NK (**Supplementary Table 5**; Methods: COVID-19 PBMC scATACseq data analysis). This produced three validation peak sets for mild COVID-19 infection. For severe COVID-19, an existing study focused on T cells identified specific chromatin activity changes with severe COVID-19 in CD8 T cells^27^. We used their reported chromatin sites for validating the circuit chromatin sites identified in CD8 T cells. In all four validation sets, the precision (proportion of sites that are differential in the validation data) of the MAGICAL-selected chromatin sites is significantly higher than the original DAS (*p*-values <0.001, two-side Fisher’s exact test, **Figures 2c** and **2d**).

When multiple potential chromatin regulatory loci are identified in the vicinity of a specific gene, it is commonly assumed that the locus closest to the transcriptional starting site (TSS) is likely to be the most important regulatory site. Challenging this assumption, however, are the results of experimental studies showing that genes may not be regulated by the nearest region^28, 29^. Supporting the importance of more distal regulatory loci, MAGICAL-selected chromatin sites significantly outperformed the nearest DAS to the TSS of DEG or all DAS within the same TAD with DEG, and the improvement is substantial (precision is ∼50% better with MAGICAL, *p-*values<0.05, two-side Fisher’s exact test, **Figures 2c** and **2d**).

To validate the circuit genes modulated by mild or severe COVID-19, we used genes reported by external COVID-19 single-cell studies^21, 30, 31^. In total, we collected six validation gene sets (three cell types for mild COVID-19 and three cell types for severe COVID-19). The precision of MAGICAL-selected circuit genes is significantly higher than that of original DEG in all validations (precision is ∼30% better with MAGICAL, *p-* values<0.05, two-side Fisher’s exact test, **Figures 2e** and **2f**). These results confirmed the increased accuracy of disease association for both chromatin sites and genes in MAGICAL-identified regulatory circuits.

### MAGICAL analysis of *S. aureus* single-cell multiomics data

We applied MAGICAL to the clinically important challenge of distinguishing methicillin-resistant (MRSA) and methicillin-susceptible *S. aureus* (MSSA) infections^32–34^. We profiled paired scRNA-seq and scATAC-seq data using human PBMCs from adults who were blood culture positive for *S. aureus*, including 10 MRSA and 11 MSSA, and from 23 uninfected control subjects (**Figure 3a; Supplementary Table 6**). To integrate scRNA-seq data from all samples, we implemented a Seurat^35^-based batch correction and cell type annotation pipeline (Methods: *S. aureus* scRNA-seq data analysis). In total, 276,200 quality cells were selected and labeled (**Figure 3b; Supplementary Figure 4; Supplementary Table 7**). For scATAC-seq data, we integrated the fragment files from quality samples using ArchR^25^ and selected and annotated 70,174 quality cells (**Figure 3c; Supplementary Figure 5; Supplementary Table 8**). In total, 388,860 peaks were identified (**Supplementary Figure 5b; Supplementary Table 9**; Methods: *S. aureus* scATAC-seq data analysis). 13 major cell types that surpassed the 200-cell threshold in both scRNA-seq and scATAC-seq data were selected for subsequent analysis (**Supplementary Figure 6**). Differential analysis for three contrasts (MRSA vs Control, MSSA vs Control, and MRSA vs MSSA) in each cell type returned a total of 1,477 DEG and 23,434 DAS (**Supplementary Figure 7; Supplementary Tables 10 and 11**).

**Figure 3.**
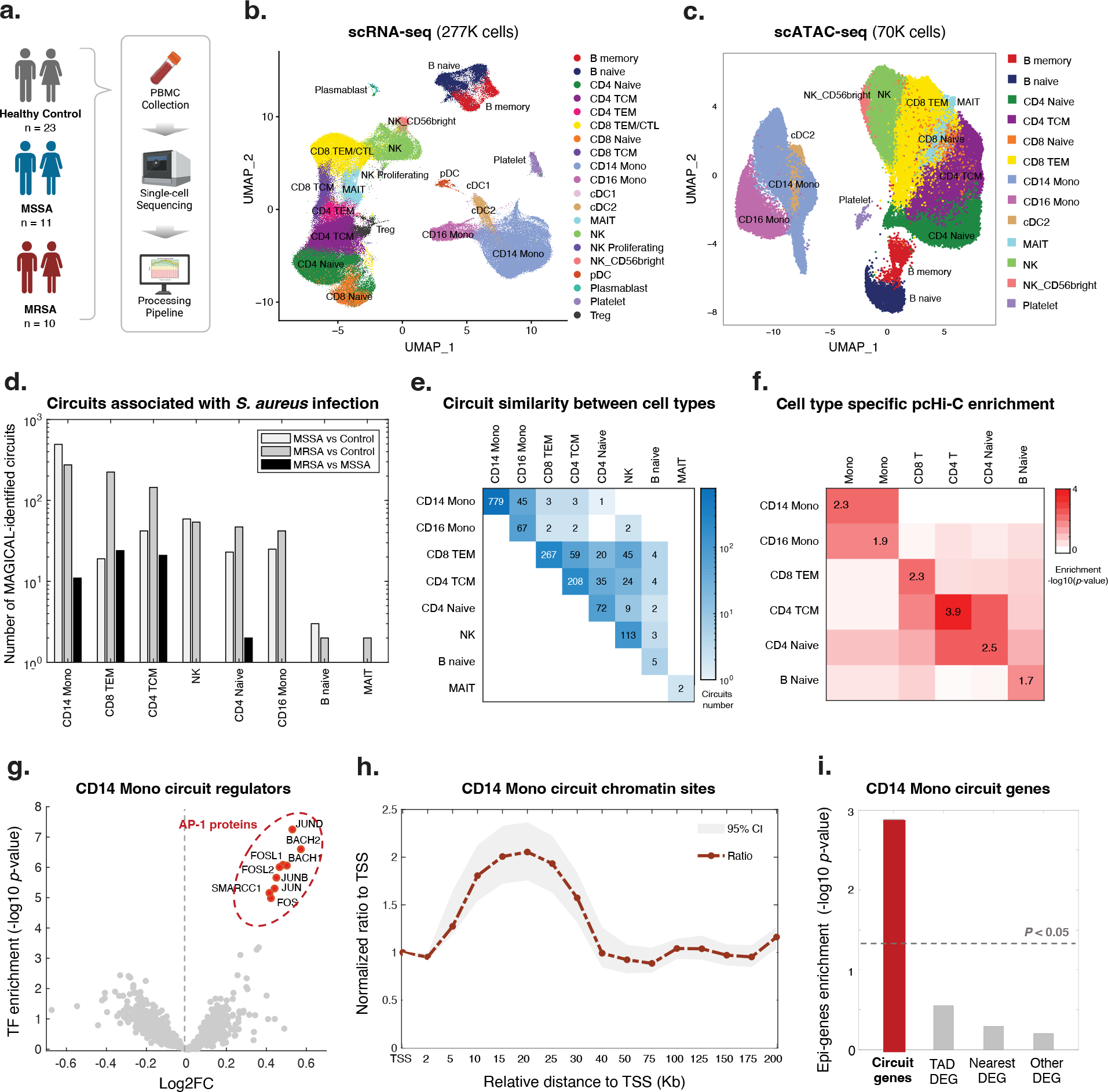
MAGICAL accurately identified distal regulatory chromatin sites and epi-driven genes associated with *S. aureus* infection. (**a**) We collected PBMC samples from 10 MRSA-infected, 11 MSSA-infected, and 23 healthy control subjects and generated same-sample scRNA-seq and scATAC-seq data using separate assays. (**b**) UMAP of integrated scRNA-seq data with 18 PBMC cell subtypes. (**c**) UMAP of integrated scATAC-seq data with 13 PBMC cell subtypes. Under-represented subtypes including cDC1, CD4 TEM, CD8 CTL, pDC, and Plasmablast, altogether representing less than 5% of cells in the scRNA-seq data, were not recovered from the scATAC-seq data. (**d**) The number of MAGICAL-identified regulatory circuits for each cell type and in contrast analysis. (**e**) The number of shared and specific circuits between cell types. (**f**) Enrichment of circuit peak-gene interactions in each cell type with cell type-specific pcHi-C interactions. (**g-i**) We specifically analyzed MAGICAL-identified regulatory circuits for CD14 monocytes. (**g**) TF motif enrichment analysis in circuit sites showed that AP-1 proteins are mostly significantly enriched at chromatin regions with increased accessibility in the infection condition. The log2FC is calculated for each TF by dividing the number of binding sites with increased chromatin activity in the infection condition by the number of sites with decreased activity. (**h**) In total, 633 circuit sites were identified by MAGICAL. In comparison to all accessible chromatin sites, an increased proportion of circuit sites were in the range of 15Kb to 25Kb relative to gene TSS. The center points represent the fold change between the proportion of circuit sites and background sites in each window. The upper and lower points represent the 95% confidence interval. (**i**) The circuit genes were significantly enriched with experimentally confirmed epi-genes in monocytes. All significance evaluation is assessed using the adjusted *p*-value of one-side hypergeometric test.

MAGICAL identified 1,513 high-confidence regulatory circuits (1,179 sites and 371 genes) within cell types for three contrasts (MRSA vs Control, MSSA vs Control, and MRSA vs MSSA) (**Supplementary Table 12**; Methods: MAGICAL analysis of *S.aureus* single-cell multiomics data). It has been reported that activation of CD14 monocytes plays a principal role in response to *S. aureus* infection^36, 37^. In MAGICAL analysis, CD14 monocytes showed the highest number of regulatory circuits (**Figure 3d**). Comparing circuits between cell types we found that these disease-associated circuits are cell type-specific (**Figure 3e**). For example, circuits rarely overlapped between very distinct cell types like monocytes and T cells. Between CD14 mono and CD16 mono, or between subtypes of T cells, most circuits are still specific for one cell type. These circuits were further validated using cell type-specific chromatin interactions reported in a reference promoter capture (pc) Hi-C dataset^13^. In all the cell types for which the cell type-specific pcHi-C data was available (B cells, CD4 T cells, CD8 T cells, CD14 monocytes), the circuit peak-gene interactions showed significant enrichment of pcHi-C interactions in matched cell types (**Figure 3f**; *p*-values < 0.01, one-side hypergeometric test). For comparison, we also performed the peak-gene interaction enrichment analysis between different cell types, finding significantly lower enrichment levels. These results indicate cell-type specificity of MAGICAL-identified circuits.

In CD14 monocytes, MAGICAL identified AP-1 complex proteins as the most important regulators, especially at chromatin sites showing increased activity in infection cells (**Figure 3g**). This finding is consistent with the importance of these complexes in gene regulation in response to a variety of infections^5, 38, 39^. Supporting the accuracy of the identified TFs, we compared circuit chromatin sites with ChIP-seq peaks from the Cistrome database^40^. The most similar TF ChIP-seq profiles were from AP-1 complex JUN/FOS proteins in blood or bone marrow samples (**Supplementary Figure 8**). Moreover, functional enrichment analysis^41^ of the circuit genes showed that cytokine signaling, a known pathway mediated by AP-1 factors and associated with the inflammatory responses in macrophages^42, 43^, was the most enriched (adjusted *p*-value 2.4e-11, one-side hypergeometric test).

MAGICAL modeled regulatory effects of both proximal and distal regions on genes. We examined the chromatin site location relative to the target gene TSS, for circuits chromatin sites and genes identified for CD14 monocytes. Compared to all ATAC peaks called around the circuit genes, a substantially increased proportion of circuit chromatin sites were located 15Kb to 25Kb away from the TSS (**Figure 3h**). This pattern is consistent with the 24Kb median enhancer distance found by CRISPR-based perturbation in a blood cell line^44^. In addition, nearly 50% of circuit chromatin sites were overlapping with enhancer-like regions in the ENCODE database^45^, further emphasizing that MAGICAL circuits are enriched in distal regulatory loci. We also found that these circuit chromatin sites were significantly enriched in inflammatory-associated genomic loci reported in the genome-wide association studies (GWAS) catalog database^46^, suggesting active host epigenetic responses to infectious diseases (**Supplementary Figure 9**; *p*-value < 0.005 when compared to control diseases, two-wide Wilcoxon rank sum test). Notably, one distal chromatin site (hg38 chr6: 32,484,007-32,484,507) looping to HLA-DRB1 is within the most significant GWAS region (hg38 chr6: 32,431,410-32,576,834) associated with *S. aureus* infection^47^.

We finally compared circuit genes to existing epi-genes whose transcriptions were significantly driven by epigenetic perturbations in CD14 monocytes^48^. MAGICAL-identified circuit genes were significantly enriched with epi-genes (**Figure 3i**; adjusted *p*- value < 0.005, one-side hypergeometric test) while the remaining DEG not selected by MAGICAL, or those mappable with DAS either within the same topological domains or closest to each other showed no evidence of being epigenetically driven. These results suggest that MAGICAL accurately identified regulatory circuits activated in response to *S. aureus* infection.

### S. aureus infection prediction

Early diagnosis of *S. aureus* infection and the strain antibiotic sensitivity is critical to appropriate treatment for this life-threatening condition. We first evaluated whether the MAGIC-identified circuit genes that are in common to MRSA and MSSA could provide a robust signature for predicting the diagnosis of *S. aureus* infection in general. Within each cell type, we selected circuit genes common to both the MRSA and MSSA analyses, resulting in 152 genes (**Figure 4a; Supplementary Table 12**). To evaluate this *S. aureus* infection, we collected external, public expression data of *S. aureus* infected subjects. In total, we found one adult whole-blood^49^ and two pediatric PBMC bulk microarray datasets^50, 51^ that comprised a total of 126 *S. aureus* infected subjects and 68 uninfected controls. The use of pediatric validation data has the advantage of providing a much more rigorous test of the robustness of MAGICAL-identified circuit genes for classifying disease samples in this very different cohort.

**Figure 4:**
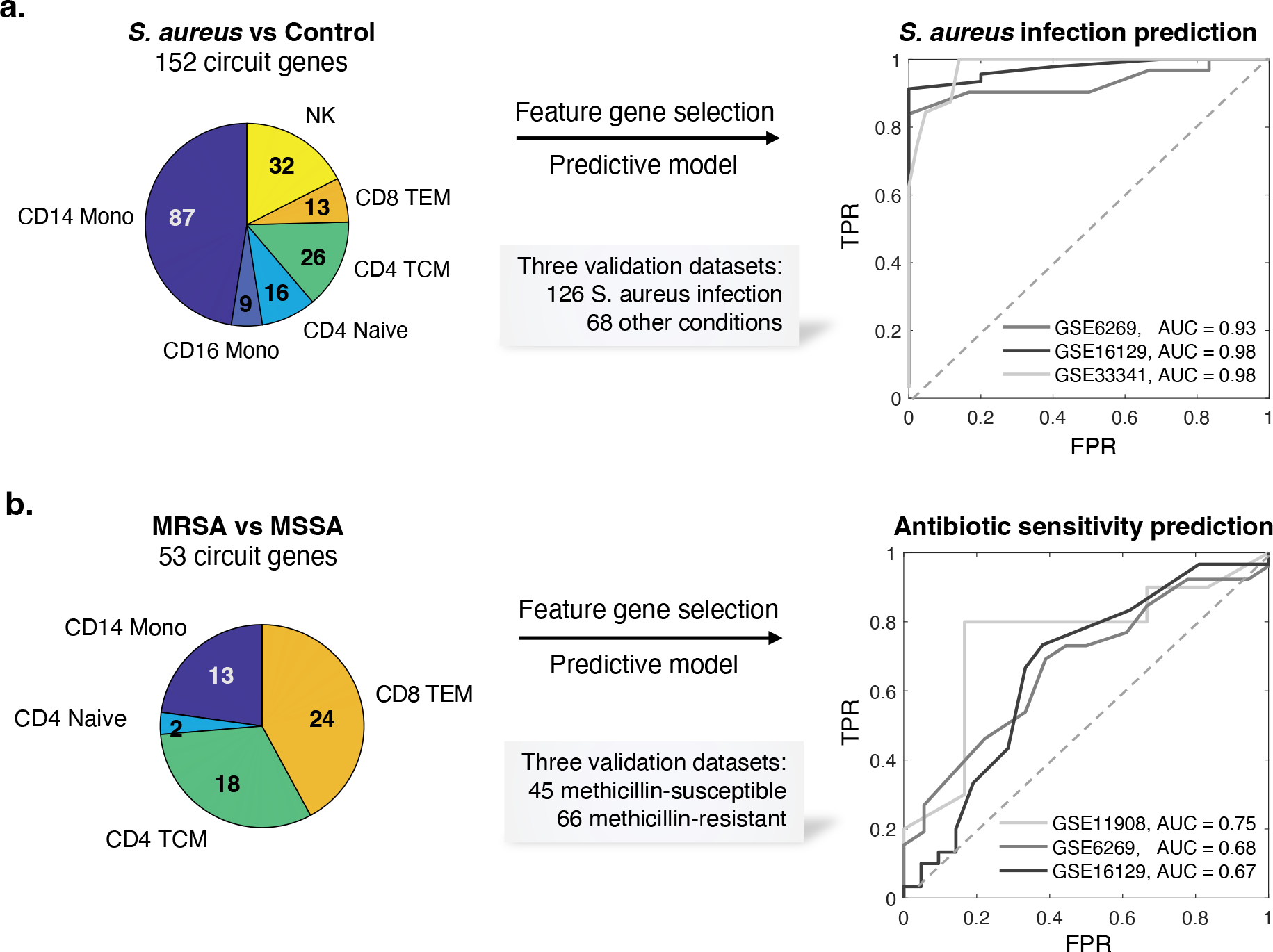
MAGICAL-identified circuit genes robustly predict *S. aureus* infection and bacteria antibody sensitivity. (**a**) Circuit genes in common to MRSA and MSSA infections achieved a near-perfect classification of *S. aureus* infected and uninfected samples in multiple independent datasets (one adult dataset and two pediatric datasets). (**b**) Circuit genes that differed between MRSA and MSSA showed predictive value of antibiotic sensitivity in independent patient samples (three pediatric datasets).

To allow validation using public bulk transcriptome datasets, we refined the 152 circuit genes set by selecting those with robust performance in our dataset at pseudobulk level. We calculated an AUROC for each circuit gene by classifying *S. aureus* infection and control subjects using pseudobulk gene expression (aggregated from the discovery scRNA-seq data). 117 circuit genes with AUROCs greater than 0.7 were selected (**Supplementary Table 13; Supplementary Figure 10a**). Functional gene enrichment analysis showed that IL-17 signaling was significantly enriched (adjusted *p*-value 2.4e- 4, **one-side hypergeometric test**), including genes from AP-1, Hsp90, and S100 families. IL-17 had been found to be essential for the host defense against cutaneous *S. aureus* infection in mouse models^52^. We trained an SVM model using the selected circuit genes as features and the discovery pseudobulk gene expression data as input. We then applied the trained SVM model to each of the three validation datasets. The model achieved high prediction performance on all datasets, showing AUROCs from 0.93 to 0.98 (**Figure 4a**).

This generalizability of circuit genes for predicting infection in different cohorts suggested that MAGICAL identifies regulatory processes that are fundamental to the host response to *S. aureus* sepsis. We further evaluated this by comparing the 117 circuit genes to 366 filtered DEG (with per gene AUROC>0.7 in the discovery pseudobulk gene expression data). We examined the differential expression π-value^53^ (a statistic score that combines both fold change and p-values) of genes in the validation datasets and found significantly higher π-values for the circuit genes (**Supplementary Figure 10b**; *p*-value 9.0e-3, one-side Wilcoxon rank sum test).

### *S. aureus* antibiotic sensitivity prediction

We then addressed the challenging problem of predicting strain antibiotic sensitivity in *S. aureus* infection. When we tested the predictive models trained with DEG for the contrast of MRSA and MSSA on three pediatric PBMC microarray datasets^50, 51, 54^ (comprising a total of 66 MRSA and 45 MSSA samples), we did not find predictive value (median of prediction AUCs close to 0.5) (**Supplementary Figure 10d-f**). And in all tests, the statistical difference between DEG-based prediction scores of the MRSA and MSSA samples in the validation datasets was never significant. These results suggest that using host scRNA-seq data alone fails to identify robust features for predicting the antibiotic sensitivity of the infected strain. These echo previous studies showing that in challenging cases, differential expression analysis using RNA-seq data had limited power to identify robust features for disease-control sample classification^55^.

With MAGICAL we identified 53 circuit genes from the comparative multiomics data analysis between MRSA and MSSA (**Supplementary Table 14**). A model trained using 32 circuit genes that were robustly differential in the discovery pseudobulk data (per gene discovery AUROC > 0.7, **Supplementary Figure 10c**) best distinguished antibiotic-resistant and antibiotic-sensitive samples in all three validation datasets, with AUROCs from 0.67 to 0.75 (**Figure 4b**). And the statistical difference between prediction scores of MRSA and MSSA samples was significant (p-value = 9.2e-3, **two-side Wilcoxon rank sum test**). The success of the circuit gene-based model demonstrated that MAGICAL captured generalizable regulatory differences in the host immune response to these closely related bacterial infections.

## Discussion

MAGICAL addressed the previously unmet need of identifying differential regulatory circuits based on single cell multiomics data from different conditions. Critically, it identifies regulatory circuits involving distal chromatin sites. The previously difficult-to-predict distal regulatory regions are increasingly recognized as key for understanding gene regulatory mechanisms. As MAGICAL uses DAS and DEG called from a pre-selected cell type, for less distinct cell types or conditions, it is harder for MAGICAL to infer circuits at cell type resolution as there will be few candidate peaks and genes. Also, MAGICAL analyzes each cell type separately, and cell type specificity is not directly modeled for disease circuit identification. Incorporating an approach to directly identify cell type-specific circuits regulated in disease conditions would be valuable. In future work, we hope to extend the MAGICAL framework to improve circuit identification when cell types are poorly defined and to model cell type specificity.

## Methods

### Human participants

The COVID-19 study protocol was approved by the Naval Medical Research Center institutional review board (protocol number NMRC.2020.0006) in compliance with all applicable Federal regulations governing the protection of human subjects. The staphylococcus sepsis protocol was reviewed and approved by the Duke Medical School institutional review board (protocol number Pro00102421). Subjects provided written informed consent prior to participation.

### Statistics & Reproducibility

No statistical methods were used to pre-determine sample sizes. No data were excluded from the analyses. The experiments were not randomized. The Investigators were not blinded to allocation during experiments and outcome assessment.

### *S. aureus* patient and control samples selection

Patients with culture-confirmed *S. aureus* bloodstream infection transferred to DUMC are eligible if pathogen speciation and antibiotic susceptibilities are confirmed by the Duke Clinical Microbiology Laboratory. DNA and RNA samples, PBMCs, clinical data, and the bacterial isolate from the subject are cataloged using an IRB-approved Notification of Decedent Research. We excluded samples if prior enrollment of the patient in this investigation (to ensure statistical independence of observations) or they are polymicrobial (i.e., more than one organism in blood or urine culture). In total, 21 adult patients were selected with 10 MRSAs and 11 MSSAs. None of them received any antibiotics in the 24 h before the bloodstream infection. Control samples were obtained from uninfected healthy adults matching the sample number and age range of the patient group. In total, 23 samples were collected from two cohorts: 14 controls provided by from the Weill Cornell Medicine, New York, NY, and 9 controls (provided by the Battelle Memorial Institute, Columbus, OH. Meta information of the selected subjects were provided in **Supplementary Table 6**.

### PBMC thawing

Frozen PBMC vials were thawed in a 37°C-waterbath for 1 to 2 minutes and placed on ice. 500μl of RPMI/20% FBS was added dropwise to the thawed vial, the content was aspirated and added dropwise to 9ml of RPMI/20% FBS. The tube was gently inverted to mix, before being centrifuged at 300xg for 5 min. After removal of the supernatant, the pellet was resuspended in 1-5ml of RPMI/10% FBS depending on the size of the pellet. Cell count and viability were assessed with Trypan Blue on a Countess II cell counter (Invitrogen).

### *S. aureus* scRNA-seq data generation

ScRNA-seq was performed as described (10x Genomics, Pleasanton, CA), following the Single Cell 3’ Reagents Kits V3.1 User Guidelines. Cells were filtered, counted on a Countess instrument, and resuspended at a concentration of 1,000 cells/μl. The number of cells loaded on the chip was determined based on the 10X Genomics protocol. The 10X chip (Chromium Single Cell 3’ Chip kit G PN-200177) was loaded to target 5,000- 10,000 cells final. Reverse transcription was performed in the emulsion and cDNA was amplified following the Chromium protocol. Quality control and quantification of the amplified cDNA were assessed on a Bioanalyzer (High-Sensitivity DNA Bioanalyzer kit) and the library was constructed. Each library was tagged with a different index for multiplexing (Chromium i7 Multiplex Single Index Plate T Set A, PN-2000240) and quality controlled by Bioanalyzer prior to sequencing.

### *S. aureus* scRNA-seq data analysis

Reads of scRNA-seq experiments were aligned to human reference genome (hg38) using 10x Genomics Cell Ranger software (version 1.2). The filtered feature-by-barcode count matrices were then processed using Seurat^35^. Quality cells were selected as those with more than 400 features (transcripts), fewer than 5,000 features, and less than 10% of mitochondrial content (**Supplementary Figure 4; Supplementary Table 7**). Cell cycle phase scores were calculated using the canonical markers for G2M and S phases embedded in the Seurat package. Finally, the effects of mitochondrial reads and cell cycle heterogeneity were regressed out using SCTransform.

To integrate cells from heterogeneous disease samples, we first built a reference by integrating and annotating cells from the uninfected control samples using a Seurat-based pipeline. For batch correction, we identified the intrinsic batch variants and used Seurat to integrate cells together with the inferred batch labels. All control samples were integrated into one harmonized query matrix. Each cell was assigned a cell type label by referring to a reference PBMC single cell dataset. The cell type label of each cell cluster was determined by most cell labels in each. Canonical markers were used to refine the cell type label assignment. This integrated control object was used as reference to map the infected samples.

To avoid artificially removing the biological variance between each infected sample during batch correction, we computationally predicted and manually refined cell types for each sample. All infection samples were projected onto the UMAP of the control object for visualization purpose. In total, 276,200 high-quality cells and 19 cell types with at least 200 cells in each were selected for the subsequent analysis. Within each cell type, differentially expressed genes (DEG) between contrast conditions were first called using the “Findmarkers” function of the Seurat V4 package^35^ with default parameters. DEG with Wilcoxon test FDR < 0.05, |log2FC|>0.1 and actively expressed in at least 10% cells (pct>0.1) from either condition were selected. To correct potential bias caused by the different sequencing depth between samples, we ran DEseq2^56^ on the aggregated pseudobulk gene expression data. Refined DEG passing pseudobulk differential statistics *p*-value <0.05 and |log2FC|>0.3 were selected as the final DEG (**Supplementary Table 10**).

### Nuclei isolation for scATACseq

Thawed PBMCs were washed with PBS/0.04% BSA. Cells were counted and 100,000- 1,000,000 cells were added to a 2mL-microcentrifuge tube. Cells were centrifuged at 300xg for 5min at 4°C. The supernatant carefully completely removed, and 0.1X lysis buffer (1x: 10mM Tris-HCl pH 7.5, 10mM NaCl, 3mM MgCl2, nuclease-free H2O, 0.1% v/v NP-40, 0.1% v/v Tween-20, 0.01% v/v digitonin) was added. After 3min incubation on ice, 1ml of chilled wash buffer was added. The nuclei were pelted at 500xg for 5min at 4°C and resuspended in a chilled diluted nuclei buffer (10X Genomics) for scATAC-seq. Nuclei were counted and the concentration was adjusted to run the assay.

### *S. aureus* scATAC-seq data generation

ScATAC-seq was performed immediately after nuclei isolation and following the Chromium Single Cell ATAC Reagent Kits V1.1 User Guide (10x Genomics, Pleasanton, CA). Transposition was performed in 10μl at 37°C for 60min on at least 1,000 nuclei, before loading of the Chromium Chip H (PN-2000180). Barcoding was performed in the emulsion (12 cycles) following the Chromium protocol. After post GEM cleanup, libraries were prepared following the protocol and were indexed for multiplexing (Chromium i7 Sample Index N, Set A kit PN-3000427). Each library was assessed on a Bioanalyzer (High-Sensitivity DNA Bioanalyzer kit).

### *S. aureus* scATAC-seq data analysis

Reads of scATAC-seq experiments were aligned to human reference genome (hg38) using 10x Genomics Cell Ranger software (version 1.2). The resulting fragment files were processed using ArchR^25^. Quality cells were selected as those with TSS enrichment > 12, the number of fragments >3000 and <30000, and nucleosome ratio <2 (**Supplementary Figure 5a; Supplementary Table 8**). The likelihood of doublet cells was computationally assessed using ArchR’s addDoubletScores function and cells were filtered using the ArchR’s filterDoublets function with default settings. Cells passing quality and doublet filters from each sample were combined into a linear dimensionality reduction using ArchR’s addIterativeLSI function with the input of the tile matrix (read counts in binned 500bps across the whole genome) with iterations = 2 and varFeatures = 20000. This dimensionality reduction was then corrected for batch effect using the Harmony method^57^, via ArchR’s addHarmony function. The cells were then clustered based on the batch-corrected dimensions using ArchR’s addClusters function. We annotated scATAC-seq cells using ArchR’s addGeneIntegrationMatrix function, referring to a labeled multimodal PBMC single cell dataset. Doublet clusters containing a mixture of many cell types were manually identified and removed. In total, 70,174 high-quality cells and 13 cell types with at least 200 cells in each were selected.

Peaks were called for each cell type using ArchR’s addReproduciblePeakSet function with the MACS2 peak caller^26^ (**Supplementary Figure 5b**). In total, 388,859 peaks were identified (**Supplementary Table 9**). Within each cell type, differentially accessible chromatin sites (DAS) between contrast conditions (MRSA vs Control, MSSA vs Control or MRSA vs MSSA) were called from the single cell chromatin accessibility count data using the “getMarkerFeatures” function of ArchR v1.0.2^25^, with parameter settings as testMethod = "wilcoxon", bias = "log10(nFrags)", normBy = "ReadsInPeaks", and maxCells = 15000. Peaks with single cell differential statistics FDR < 0.05, |log2FC|>0.1, and actively accessible in at least 10% cells (pct>0.1) from either condition were selected as DAS. Due to the high false positive rate in single cell-based differential analysis^58^, we further refined the DAS by fitting a linear model to the aggregated and normalized pseudobulk chromatin accessibility data and tested DAS individually about their covariance with sample conditions^56^. Refined DAS passing pseudobulk differential statistics *p*-value <0.05 and |log2FC|>0.3 between the contrast conditions were selected as the final DAS (**Supplementary Table 11**).

### MAGICAL

To build candidate regulatory circuits, TFs were mapped to the selected DAS by searching for human TF motifs from the chromVARmotifs library^59^ using ArchR’s addMotifAnnotations function. The binding DAS were then linked with DEG by requiring them in the same TAD within boundaries. Then, a candidate circuit is constructed with a chromatin region and a gene in the same domain, with at least one TF motif match in the region.

For each cell type (i.e. *i*-th cell type), MAGICAL inferred the confidence of TF-peak binding and peak-gene looping in each candidate circuit using a hierarchical Bayesian framework with two models: a model of TF-peak binding confidence (***B***) and hidden TF activity (***T***) to fit chromatin accessibility (***A***) for *M* TFs and *P* chromatin sites in *K_A_*_,*S*_, *i* cells with scATAC-seq measures from *S* samples; a second model of peak-gene interaction (***L***) and the refined (noise removed) regulatory region activity (***BT***) to fit gene expression (***R***) of *G* genes in *K_R_*_,*S*_, *i* cells with scRNA-seq measures from the same *S* samples.

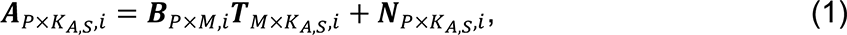

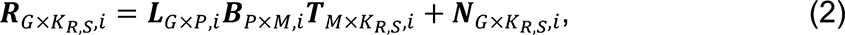

***A***_*P*×*K*_*A*,*S*__,*i* was a *P* by *K_A_*_,*S*_, *i* matrix with each element *a*_*p*,*k*_*A*,*S*__,*i* representing the ATAC read count of *p*-th chromatin site (ATAC peak) in *k_A_*_,*S*_-th cell in *s*-th sample.

***R***_*G*×*K*_*R*,*S*__,*i* was a *G* by *K_R_*_,*S*_, *i* matrix with each element *r*_*g*,*k*_*R*,*s*__,*i* representing the RNA read count of *g*-th gene in *k_R_*_,*S*_-th cell of *s*-th sample.

***N***_*P*×*K*_*A*,*S*,__*i* and ***N***_*G*×*K*_*R*,*S*,*i*__ represented data noise in corresponding to ***A***_*P*×*K*_*A*,*S*,__*i* and ***R***_*G*×*K*_*R*,*S*,*i*__.

***B***_*P*×*M*,*i*_ was a *P* by *M* matrix with each element *b_p,m,i_* representing the binding confidence of *m*-th TF on *p*-th candidate chromatin site.

***L***_*G*×*P*,*i*_ was a *G* by *P* matrix with each element *l_*p*,*g*,*i*_* representing the interaction between *p*-th chromatin site and *g*-th gene.

***T***_*M*×*K*_*A*,*S*,__*i* was a *M* by *K*_*A*,*S*_,*i* matrix with each element *t*_*m*,*k*_*A*,*s*__,*i* representing the hidden TF activity of *m*-th TF in *k_A_*_,*S*_-th ATAC cell of *s*-th sample.

***T***_*M*×*K*_*R*,*S*,__*i* was a *M* by *K_R_*_,*S*_ matrix with each element *t*_*m*,*k*_*R*,*s*__,*i* representing the hidden TF activity of *m*-th TF in *k_R_*_,*S*_-th RNA cell of *s*-th sample.

***T***_*M*×*K*_*A*,*S*,__*i* and ***T***_*M*×*K*_*R*,*S*,__*i* were both extended from the same ***T****_M_*_×*S*,*i*_ (with elements *t_m_*_,*s*,*i*_) by assuming that in *i*-th cell type and *s*-th sample, *m*-th TF’s regulatory activities in all ATAC cells and all RNA cells followed an identical distribution of a single variable *t_m_*_,*s*,*i*_ Therefore, *K_A_*_,*S*,*i*_ and *K_R_*_,*S*,*i*_ can be different numbers and MAGICAL will only estimate the matrix ***T****_M_*_×*S*,*i*._

To select high-confidence regulatory circuits, MAGICAL estimated the confidence (probability) of TF-peak binding ***B****_P_*_×*M*,*i*_ and peak-gene interaction ***L****_G_*_×*P*,*i*_ together with the hidden variable ***T****_M_*_×*S*,*i*_ in a Bayesian framework.

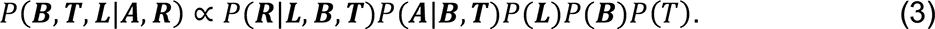

Based on the regulatory relationship among chromatin sites, upstream TFs, and downstream genes (as illustrated in **Fig. 1**), the posterior probability of each variable can be approximated as:

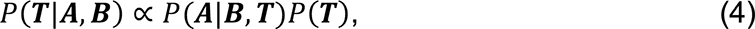

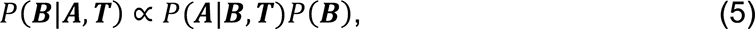

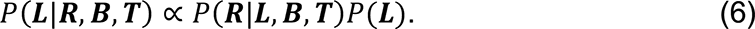

Although the prior states of *b_p_*_,*m*,*i*_ and *l_p_*_,*g*,*i*_ were obtained from the prior information of TF motif-peak mapping and topological domain-based peak-gene pairing, their values were unknown. We assumed zero-mean Gaussian priors for ***B***, ***L*** and the hidden variable ***T*** by assuming that positive regulation and negative regulation would have the same priors, which is likely to be true given the fact that there were usually similar numbers of up-regulated and down-regulated peaks and genes after the differential analysis. We set a high variance (non-informative) in each prior distribution to allow the algorithm to learn the distributions from the input data.

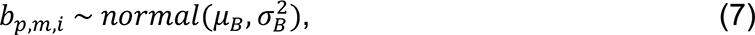

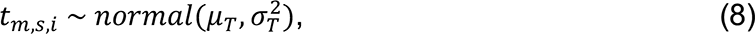

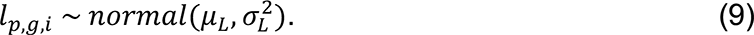

where 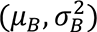, 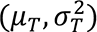, and 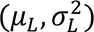 are hyperparameters representing the prior mean and variance of TF-peak binding, TF activity, and peak-gene looping variables.

The likelihood functions *P*(***A***|***B***, ***T***) and *P*(***R***|***L***, ***B***, ***T***) represent the fitting performance of the estimated variables to the input data. These two conditional probabilities are equal to the probabilities of the fitting residues ***N***_*P*×*K*_*A*,*S*,__*i* and ***N***_*G*×*K*_*R*,*S*,__*i*, for which we assumed zero-mean Gaussian distributions.

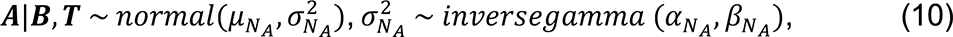

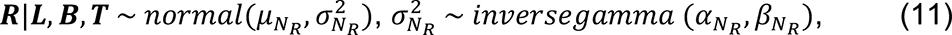

Where 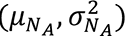 and 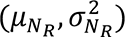 are hyperparameters representing the prior mean and variance of data noise in the ATAC and RNA measures. Here, the variance of the signal noise is modelled using inverse Gamma distributions, with hyperparameters (α_*N*_*A*__, *β*_*N*_*A*__) and (α_*N*_*R*__, *β*_*N*_*R*__) to control the variance of fitting residues (very low probabilities on large variances).

Then, the posterior probability of each variable defined in Eq. (4-6) was still a Gaussian distribution with poster mean 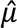 and variance 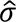 as shown below:

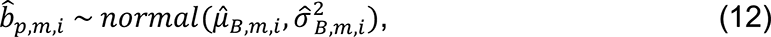

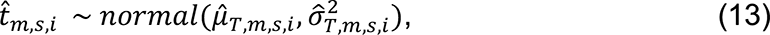

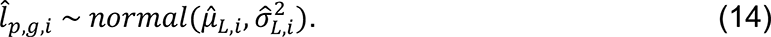

Gibbs sampling was used to iteratively learn the posterior distribution mean and variance of each set of variables and draw samples of their values accordingly.

For the TF-peak binding events, the posterior mean 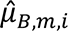 and variance 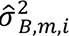 were estimated specifically for *m*-th TF since the number of binding sites and the positive or negative regulatory effects between TFs could be very different.

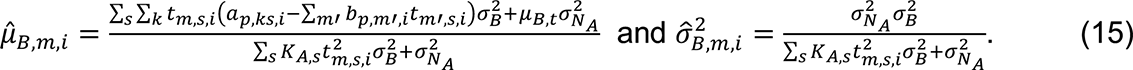

For TF activities, the posterior mean 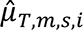 and variance 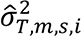 were estimated specifically for *m*-th TF and *s*-th sample using chromatin accessibility data as follows:

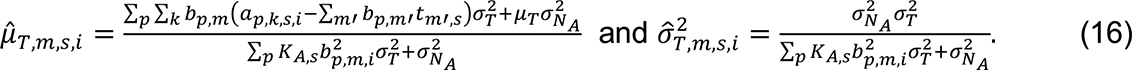

Then, based on the estimated distribution parameters of 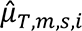 and 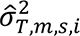 of 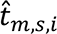, for *k_R_*_,*S*_-th RNA cell in the same *s*-th sample we draw a TF regulatory activity sample as 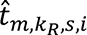 For p-th peak, we were able to reconstruct its chromatin activity in the RNA cell as 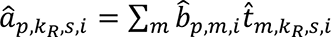, and for g-th gene, we further estimated the interaction confidence 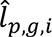 between p-th peak and g-th gene. The peak-gene interaction distribution parameters 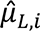 and 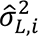 were estimated as follows:

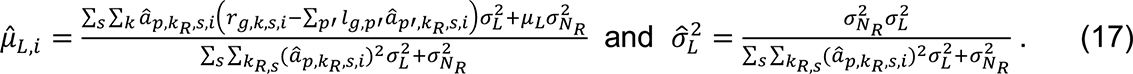

In *n*-th round of Gibbs estimation, after learning all distributions, we estimated the confidence of each linkage by linearly mapping the sampled values of 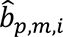 and 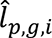 in the range of (-∞, ∞) to probabilities in (0,1) as follows:

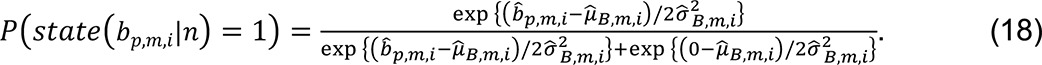

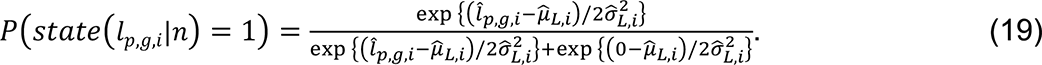

Binary state samples were then drawn based on the confidence of each linkage and were then used to initiate the next round of estimations. After running a long sampling process (in total *N* rounds) and accumulating enough samples on the binary states of TF-peak bindings and peak-gene interactions, we calculated the sampling frequency of each linkage as a posterior probability.

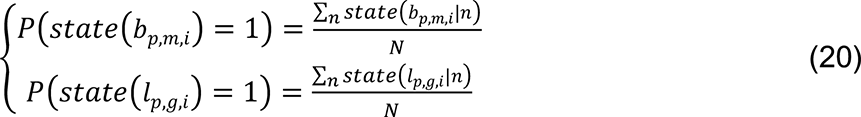

### MAGICAL analysis of *S.aureus* single-cell multiomics data

For each cell type, given DAS and DEG of contrast conditions (MRSA vs Control, MSSA vs Control or MRSA vs MSSA), MAGICAL was first initialized by mapping prior TF motifs from the ‘chromVARmotifs’ library to DAS using ArchR’s addMotifAnnotations. Because there is no PBMC cell type Hi-C data publicly available, we are using TAD boundaries from a lymphoblastoid cell line, GM12878, which was originally generated by EBV transformation of PBMCs^60^. The TAD boundary structure is closely conserved between the lymphoblastoid cell lines and primary PBMC^61^ and between cell types^62, 63^. We called TAD boundaries from a GM12878 cell line Hi-C profile^63^ using TopDom^64^. About 6000 topological domains were identified. For each contrast, we built candidate circuits by pairing DAS with TF binding sites with DEG in the same domain. MAGICAL was run 10000 times to ensure that the sampling process converged to stable states. This process was repeated for all cell types and the top 10% high confidence circuit predictions were selected from each cell type for validation analysis.

### MAGICAL analysis of COVID-19 single-cell multiomics data

As a proof of concept for contrast condition single cell multiomics data analysis, MAGICAL was applied to a public PBMC COVID-19 single-cell multiomics dataset^5^ with samples collected from patients with different severity and heathy controls. For each of the three selected cell subtypes (CD8 TEM, CD14 Mono, and NK), from the original publication we downloaded DEG for two contrasts: mild vs control and severe vs control. For each of the selected cell types, DAS were called respectively for mild vs control and severe vs control using ArchR’s functions and thresholds as introduced in the paper. MAGICAL was initialized by mapping prior TF motifs from the ‘chromVARmotifs’ library to DAS using ArchR’s addMotifAnnotations. As explained above, we used TAD boundary information of ∼6000 domains identified in GM12878 cell line^63^ as prior. Then, DAS with TF binding sites were paired with DEG in the same TAD and the initial candidate regulatory circuits were constructed. Respectively for mild and severe COVID-19, MAGICAL was run 10000 times to ensure that the sampling process converged to stable states. This process was repeated for all selected cell types. The chromatin sites and genes in the top 10% predicted high confidence circuits in each cell type were selected as disease associated.

### COVID-19 PBMC samples of validation scATAC-seq data

To validate chromatin sites associated with mild COVID-19, PBMC samples were obtained from the COVID-19 Health Action Response for Marines (CHARM) cohort study, which has been previously described^65^. The cohort is composed of Marine recruits that arrived at Marine Corps Recruit Depot—Parris Island (MCRDPI) for basic training between May and November 2020, after undergoing two quarantine periods (first a home-quarantine, and next a supervised quarantine starting at enrolment in the CHARM study) to reduce the possibility of SARS-CoV-2 infection at arrival. Participants were regularly screened for SARS-CoV-2 infection during basic training by PCR, serum samples were obtained using serum separator tubes (SST) at all visits, and a follow-up symptom questionnaire was administered. At selected visits, blood was collected in BD Vacutainer CPT Tube with Sodium Heparin and PBMC were isolated following the manufacturer’s recommendations. We used PBMC samples from six participants (five males and one female) who had a COVID-19 PCR positive test and had mild symptoms (sampled 3-11 days after the first PCR positive test), and from three control participants (three males) that had a PCR negative test at the time of sample collection and were seronegative for SARS-CoV-2 IgG. New scATAC-seq data were generated following the same protocol as described above (**Supplementary Table 2**).

### COVID-19 PBMC scATACseq data analysis

Reads of scATAC-seq experiments were aligned to human reference genome (hg38) using 10x Genomics Cell Ranger software (version 1.2). The resulting fragment files were processed using ArchR^25^. Quality cells were selected as those with TSS enrichment > 12, the number of fragments >3000 and <30000, and nucleosome ratio <2. The likelihood of doublet cells was computationally assessed using ArchR’s addDoubletScores function and cells were filtered using the ArchR’s filterDoublets function with default settings. A total of 15,836 high quality cells in the infection group and 9,125 cells in the control group were selected after QC analysis (**Supplementary Figure 3; Supplementary Table 3**). These cells were combined into a linear dimensionality reduction using ArchR’s addIterativeLSI function with the input of the tile matrix (read counts in binned 500bps across the whole genome) with iterations = 2 and varFeatures = 20000. The cells were then clustered using ArchR’s addClusters function. We annotated scATAC-seq cells using ArchR’s addGeneIntegrationMatrix function, referring to a labeled multimodal PBMC single cell dataset. Doublet clusters containing a mixture of many cell types were manually identified and removed.

Peaks were called for each cell type using ArchR’s addReproduciblePeakSet function with peak caller MACS2^26^ (**Supplementary Figure 3**). In total, 284,525 peaks were identified (**Supplementary Table 4**). For each of the three selected cell types (CD8 TEM, CD14 Mono and NK), chromatin sites with single cell differential statistics FDR < 0.05 and |log2FC|>0.1 between COVID-19 and control conditions and actively accessible in at least 10% cells (pct>0.1) from either condition were selected. Refined peaks passing pseudobulk differential statistics *p*-value <0.05 and |log2FC|>0.3 between the contrast conditions were finally selected as the validation peak set (**Supplementary Table 5**).

### COVID-19 circuit peaks and genes accuracy evaluation

The number of peaks/genes reported by each COVID-19 study would be different due to the difference in the number of recruited patients and collected cells. To overcome the issue caused by the imbalanced number between discovery and validation dataset or between differential peaks/genes and circuit sites/genes in comparison, in each comparison, the larger peak/gene set was randomly down sampled to match the smaller number of peaks/genes in the other set. The precision (site reproduction rate) is calculated to assess the accuracy of each peak/gene set.

### MAGICAL analysis of 10X PBMC single-cell true multiome data

For benchmarking, MAGICAL was applied to a 10X PBMC single cell multiome dataset including 108,377 ATAC peaks, 36,601 genes, and 11,909 cells from 14 cell types. MAGICAL used the same candidate peaks and genes as selected by TRIPOD^11^ for fair performance comparison. Two different priors were used to pair candidate peaks and genes: (1) the peaks and genes were within the same TAD from the GM12878 cell line; (2) the centers of peaks and the TSS of genes were within 500K bps. MAGICAL inferred regulatory circuits with each prior and used the top 10% predictions for accuracy assessment. High confidence peak-gene interactions predicted by TRIPOD on the same data were directly downloaded from the supplementary tables of their publication^11^. Two baseline approaches of peak-gene pairing were included: pairing all peaks with each gene if they are in the same TAD or pairing only the nearest peak to gene based on their genomic distance. To fairly assess the accuracy of MAGICAL weighted peak-gene interactions and the results (paired or non-paired) from TRIPOD or baseline approaches, we selected the top 10% predictions by MAGICAL as the final peak-gene pairing. We overlapped these pairs with the curated 3D genome interactions in blood context from the 4DGenome database^19^ and calculated the precision for each approach.

### MAGICAL analysis of GM12878 cell line SHARE-seq data

For benchmarking, MAGICAL was also applied to a GM12878 cell line SHARE-seq dataset^10^. For fair comparison, MAGICAL used the same candidate peaks and genes as selected by FigR^18^. MAGICAL was initialized with two different priors to pair candidate peaks and genes: (1) the peaks and genes were within the same prior TAD from the GM12878 cell line; (2) the centers of peaks and the TSS of genes were within 500k bps. MAGICAL inferred regulatory circuits under each setting and used the top 10% predictions for accuracy assessment. High confidence peak-gene interactions predicted by FigR were directly downloaded from the supplementary tables of the original publication^10^. Similarly, the top 10% predictions by MAGICAL and interactions paired by the two baseline approaches mentioned above were selected. We overlapped peak-gene interactions predicted by each approach with GM12878 H3K27ac HiChIP chromatin interactions^20^ for precision evaluation.

### Validating predicted peak-gene interactions

To assess the precision of the predicted circuit peak-gene interactions, we assumed a corrected inferred peak-gene pair should be also connected by a chromatin interaction reported by Hi-C or similar experiments. To check this, each peak was extended to 2kb long and then checked for overlapping with one end of a physical chromatin interaction. For genes, we checked if the gene promoter (−2kb to 500b of TSS) overlapped the other end of the interaction. Precision was calculated as the proportion of overlapped chromatin interactions among the predicted peak-gene interactions. The significance of enrichment of overlapped chromatin interactions was assessed using hypergeometric *p*- value, with all candidate peak-gene pairs as background.

### GWAS enrichment analysis

To assess the enrichment of GWAS loci of inflammatory diseases in circuit chromatin sites in each cell type, significant GWAS loci were downloaded from GWAS catalog^46^ for inflammatory diseases and control diseases. GREGOR^66^ was used to assess the enrichment of GWAS loci at which either the index SNP or at least one of its LD proxies overlaps with a circuit chromatin site, using pre-calculated LD data from 1000G EUR samples. The enrichment *p*-value of each disease GWAS was converted to a z-score. With each cell type, enrichment scores for traits with fewer than 5 overlapped GWAS SNPs with circuit sites were hold out. Also, as all reference data used by GREGOR is hg19 based, genome coordinates of testing regions were mapped from hg38 to hg19.

### Predicting *S. aureus* infection state

To refine circuit genes lately used for predicting infection diagnosis in microarray gene expression data, the capability of each circuit gene on distinguishing infection and control samples, or MRSA and MSSA samples, was assessed using sample level pseudobulk gene expression data, aggregated from the discovery scRNA-seq datasets. The total number of reads of each sample was normalized to 1e7. The normalized RNA read counts across all samples were log and z-score transformed. For each circuit gene, a discovery AUROC (area under the ROC curve) was calculated by comparing the scRNA-seq gene expression-based sample ranking against the contrasted sample groups. Circuit genes were prioritized based on AUROCs. An SVM model was trained using the top-ranked circuit genes as features and their normalized pseudobulk expression data as input. The model was then tested on independent microarray datasets. The microarray gene expression data was also log and z-score transformed to ensure a similar distribution to the training data. For comparison, top DEG prioritized by discovery AUROC or by other approaches like the Minimum Redundancy Maximum Relevance (MRMR) algorithm or LASSO regression were also tested on the same microarray datasets.

## Data availability

The 10X PBMC single cell multiome dataset can be downloaded from https://support.10xgenomics.com/single-cell-multiome-atac-gex/datasets/1.0.0/pbmc_granulocyte_sorted_10k. Users will need to provide their contact information to access the download webpage where the filtered feature barcode matrix (HDF5 format) can be downlaoded. The reference multimodal PBMC single cell dataset (H5 Seruat data file) can be downloaded from https://atlas.fredhutch.org/nygc/multimodal-pbmc/. The GWAS catalog database can be accessed at https://www.ebi.ac.uk/gwas/docs/file-downloads. SNPs associated with each disease used in this paper can be extracted from the downloadable file “All associations v1.0”. Home sapiens chromatin interactions data can be downloaded from https://4dgenome.research.chop.edu/Download.html. Home sapiens transcription factor ChIP-seq profiles can be downloaded at http://cistrome.org/db/. Users can also provide their customized peak file to the server http://dbtoolkit.cistrome.org/ and visualize transcription factors that have a significant binding overlap. Home sapiens candidate enhancers annotated by ENCODE can be downloaded at https://screen.encodeproject.org/. The chromVARmotifs library is available at https://github.com/GreenleafLab/chromVARmotifs. The source single cell data collected in this study is publicly accessible at the GEO repository (https://www.ncbi.nlm.nih.gov/geo/, accession number GSE220190) and the Zenodo repository^67–69^. The S.aureus infection scATAC-seq dataset can be accessed under subseries number GSE220188. The paired scRNA-seq dataset can be accessed under subseries number GSE220189. The COVID-19 scATAC-seq dataset can be accessed under subseries number GSE222548. Source data for Figures 2-4 is available with this manuscript.

## Code availability

The source code of MAGICAL is available on GitHub at https://github.com/xichensf/magical and the Zenodo repository^70^.

## Data Availability

Data from scRNA-seq and scATAC-seq have been deposited with the GEO repository under accession number GSE220190. All the other data used in this study are publicly available and the URLs are provided in the Data Availability section.

https://github.com/xichensf/magical

https://www.ncbi.nlm.nih.gov/geo/query/acc.cgi?acc=GSE220190

## Acknowledgments

We thank the Single-cell and Spatial Technologies team at the Center for Advanced Genomics Technology, Department of Genetics and Genomic Sciences, the Icahn School of Medicine at Mount Sinai for providing the experimental, computational, data resources, and staff expertise. S.C.S was supported by the Defense Advanced Research Projects Agency under contract N6600119C4022. A.G.L. is supported by Defense Health Agency grant 9700130 through the Naval Medical Research Center. O.G.T. is supported by National Institutes of Health under grant R01GM071966 and Simons Foundation under grant 395506. L.C.N. is supported by the National Institute of Mental Health, National Institute of Neurological Disorders and Stroke, National Institute of Diabetes and Digestive and Kidney Diseases, National Heart, Lung, and Blood Institute, and National Institute of Allergy and Infectious Diseases under grant UM1AI164599 and by National Institute on Drug Abuse under grants U01 DA53625 and U01DA058527.

## Author Contributions

S.C.S, O.G.T, and E.Z conceived the study and supervised the research. X.C. designed and implemented the computational framework, conducted benchmarks and case studies with Y.W., wrote the code, and set up the web access with the help of A.T. A.C. was involved in the S. aureus study. W.S.C. managed and processed single-cell sequencing data with help from A.R., G.N., and A.V. S.K. and D.C. conducted the public microarray data search. F.R.Z., V.N., M.C.G., and R.S. generated the PBMC single-cell multiomics data for the S. aureus infected and control subjects. The S. aureus patient blood samples were provided by C.W.W, V.G.F., F.R, and M.D. The control samples were provided by R.R.S. and L.C.N. I.R. and C.M.M. provided immunological interpretations of the results. I.R., A.G.L., and A.S.S. provided the validation PBMC scATAC-seq data of COVID-19 patients and uninfected controls. S.C.S, O.G.T, X.C., E.Z., A.C., and C.T. wrote the first draft of the manuscript. All authors proofread the submitted version.

