## Supplemental Figures 1-10 for "Mapping disease regulatory circuits at cell-type resolution from single-cell multiomics data"

### Supplementary Figures

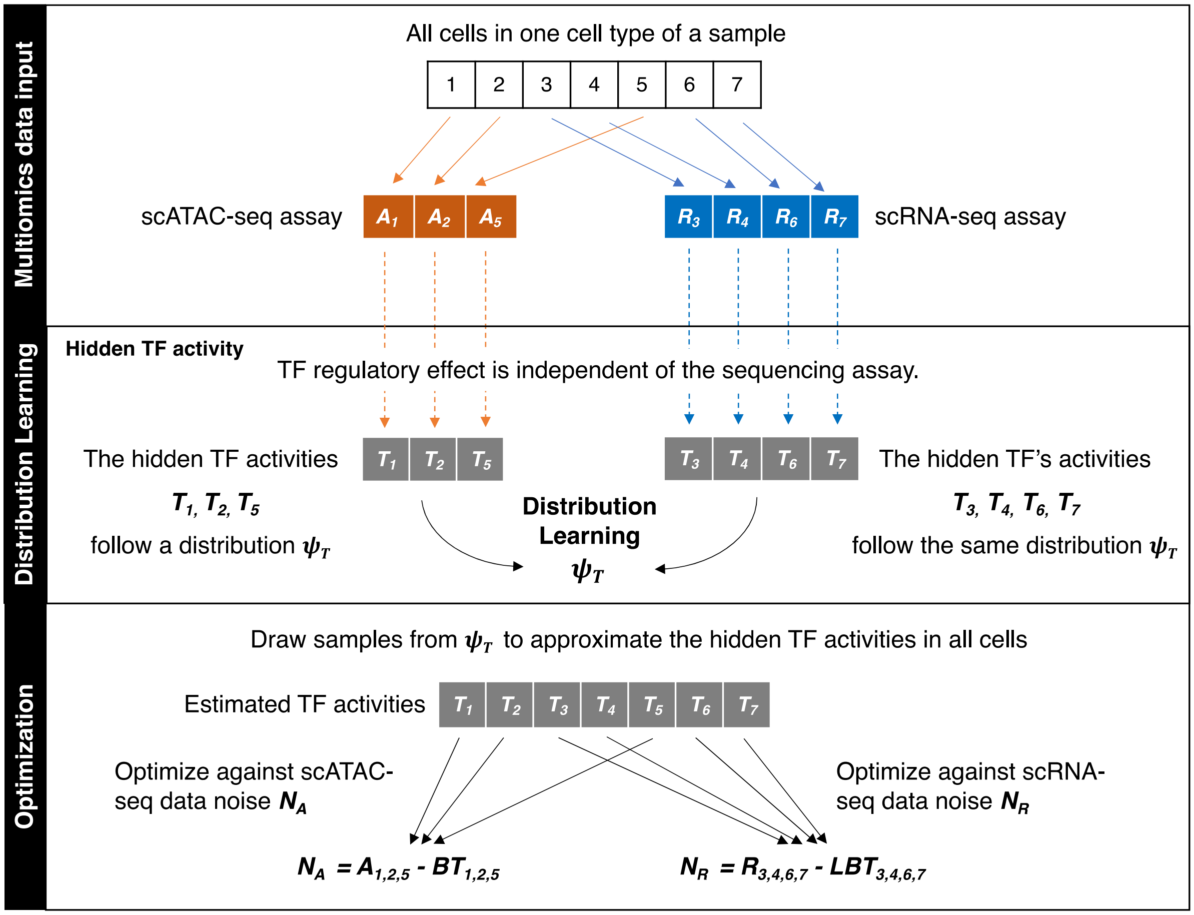


**Supplementary Figure 1. Distribution learning of the hidden TF activity.** Within one cell type of a sample, we assume that the distribution of TF activity (regulatory effect of a protein), is identical across cells from the same sample, regardless of if those cells are sequenced by the ATAC assay or RNA assay. However, there are no protein level measures so the TF activity is a hidden variable and needs to be estimated. Although precisely estimating the TF activity in each cell can be hard, its distribution can be learned from the multiomcs data. MAGICAL iteratively learns the TF activity distribution, approximates TF activities in individual cells by drawing samples from the learned distribution, and fits chromatin accessibility and gene expression data respectively using the estimated TF activity and other already estimated variables to optimize against data noise in both modalities.

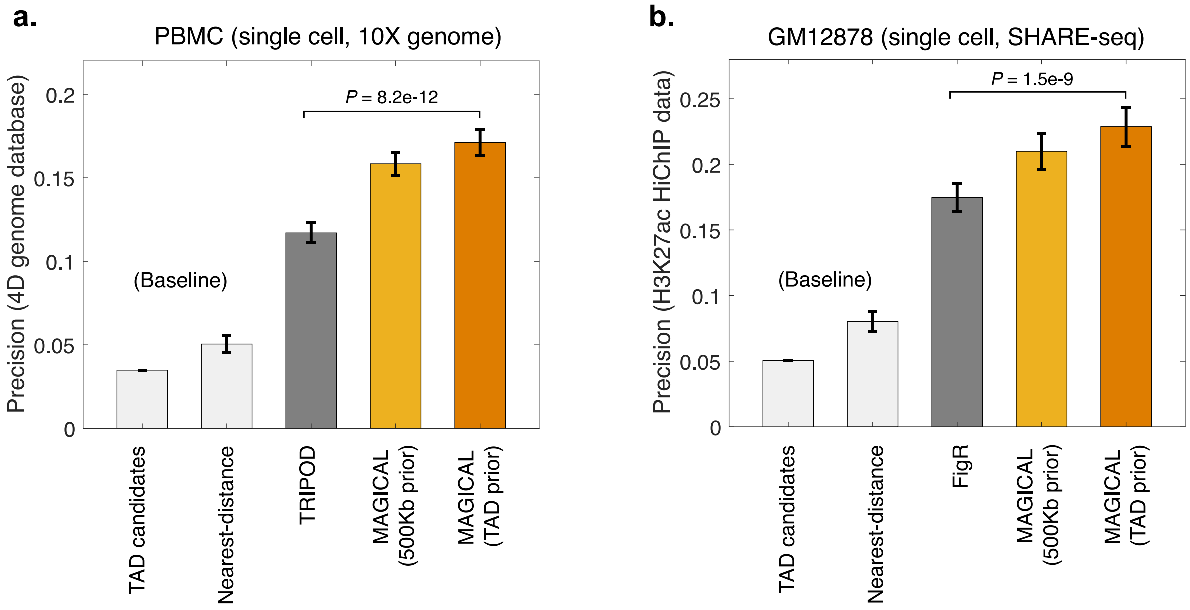


**Supplementary Figure 2. Benchmarking MAGICAL and existing methods on one condition single cell multiomics data.** (**a**) The precision of peak-gene interactions identified by each method using the 10X PBMC multiome dataset, with validation on experimental chromatin interactions in blood cells curated in the 4DGenome database. MAGICAL identified 3721 peak-gene interactions. (**b**) The precision of peak-gene interactions identified by each method using the GM12878 SHARE-seq dataset, with validation on distal chromatin interactions captured by an H3K27ac HiChIP experiment in GM12878 cell line. MAGICAL identified 5177 peak-gene interactions. Two baseline approaches are included in the comparisons as references: (1) for each candidate gene, pairing all sites with it if in the same TAD; (2) for each gene, pairing the nearest peak with it based on their genomic distance. Results were presented as boxplots where the center line represented the median of the precision after n=50 rounds of random sampling and the error bar represented the 95% confidence interval of the precision. The significance p-value was assessed using two-wide Fisher's exact test.

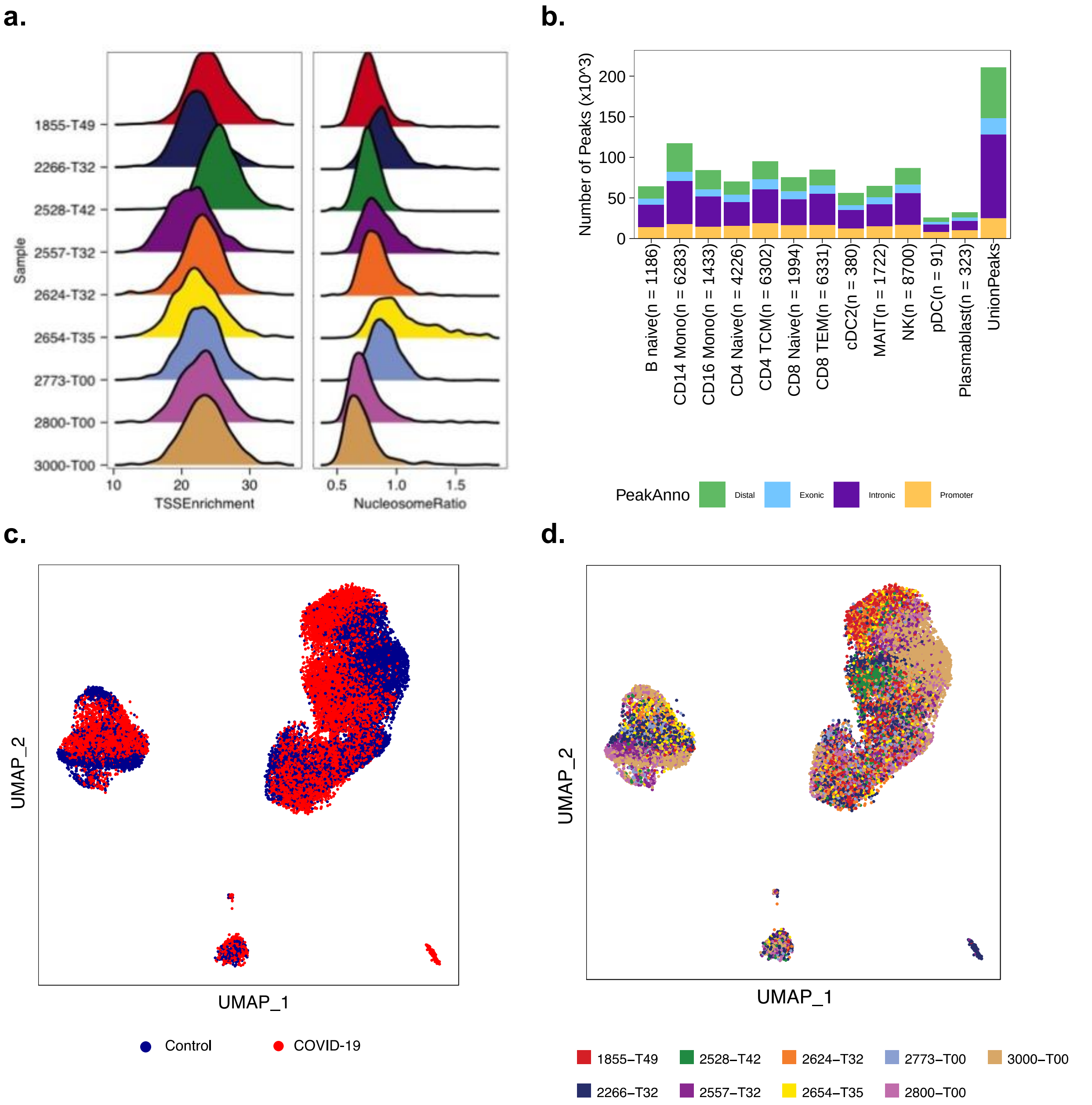


**Supplementary Figure 3. COVID-19 PBMC validation scATAC-seq data integration and peak calling using quality cells. (a)** Distribution of TSS enrichment and nucleosome ratio of cells in scATAC-seq data of 8 samples. **(b)** The number of peaks called per cell type using MACS2. Peaks are annotated as distal (>2Kb), proximal (<2Kb), exonic or intronic. **(c-d)** UMAPs of cells in the integrated scATAC-seq data with number and color representing conditions (**c**) or samples (**d**).

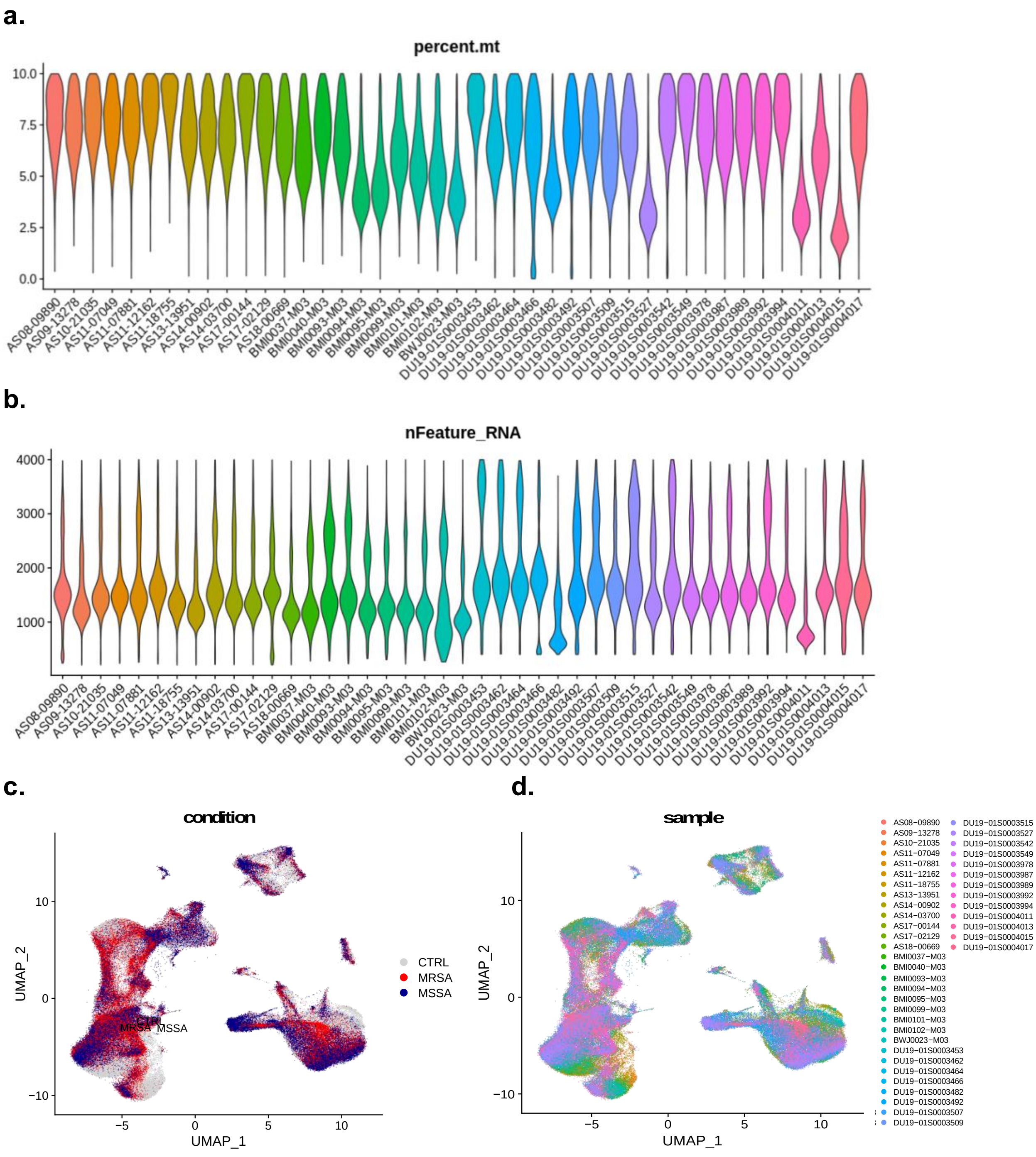


**Supplementary Figure 4.** ***S. aureus*** **PBMC scRNA-seq data integration using quality cells. (a)** Distribution of number of features (transcript) in quality cells selected for each disease sample. (**b**) Percent of mitochondrial of quality cells selected for each disease sample. **(b-c)** UMAPs of cells in the integrated object with color representing conditions or samples. Cells from all samples were well mixed in individual cell clusters, with rand index 0.016.

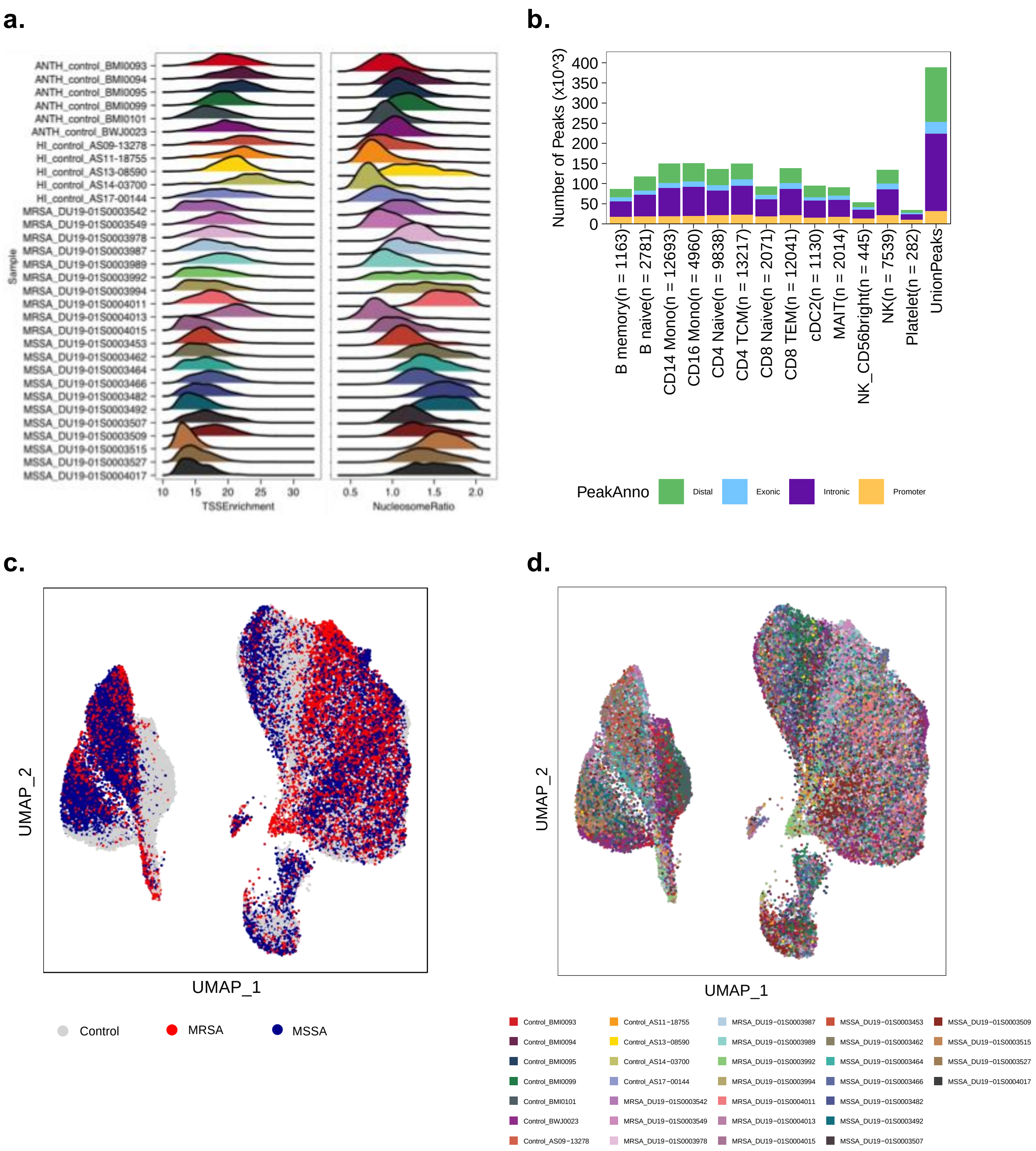


**Supplementary Figure 5.** ***S. aureus*** **PBMC scATAC-seq data integration and peak calling using quality cells. (a)** Distribution of TSS enrichment and nucleosome ratio of selected quality cells for each sample. **(b)** The number of peaks called per cell type using MACS2. Peaks are annotated as distal (>2Kb), proximal (<2Kb), exonic or intronic. **(c-d)** UMAPs of cells in the integrated scATAC-seq data with number and color representing conditions (**c**) or samples (**d**). Cells from all samples were well mixed in individual cell clusters, with rand index 0.033.

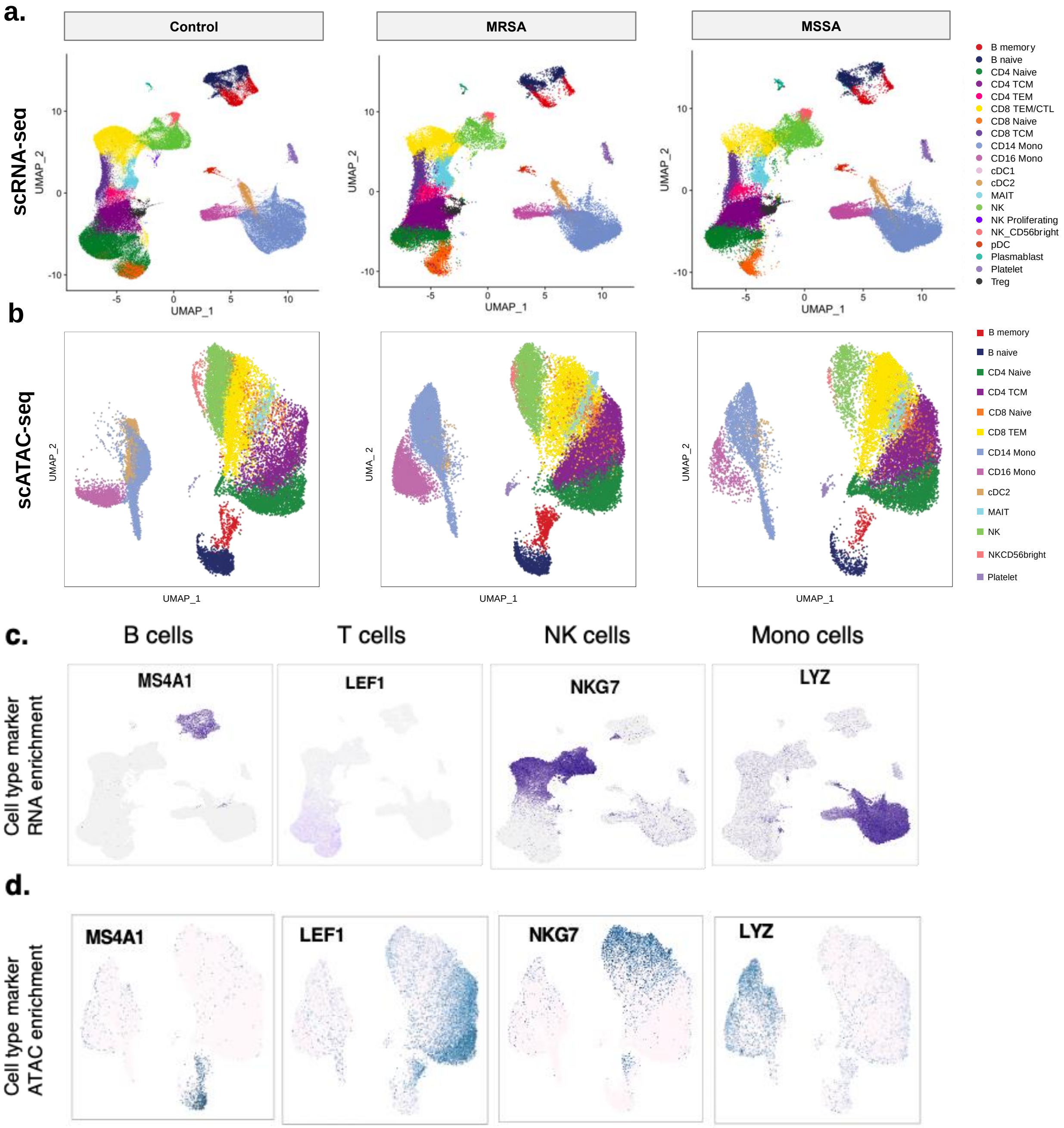


**Supplementary Figure 6. Integrated scRNA-seq and scATAC-seq data for MRSA, MSSA, and uninfected control samples.** (**a**) UMAP of scRNA-seq data for each sample group with color representing cell types. (**b**) UMAP of scATAC-seq data for each sample group with number and color representing cell types. (**c**) UMAPs of gene expression of cell type markers in the identified cell types. (**d**) UMAPs of chromatin accessibility (gene TSS + body) of cell type markers.

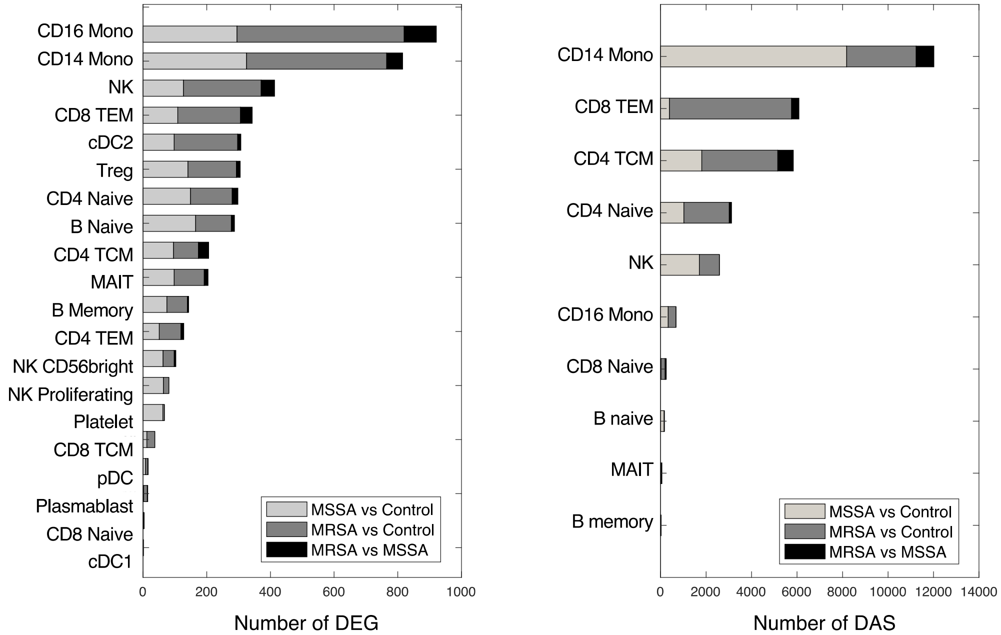


**Supplementary Figure 7. Number of DEG or DAS identified for each contrast analysis within individual cell types.**

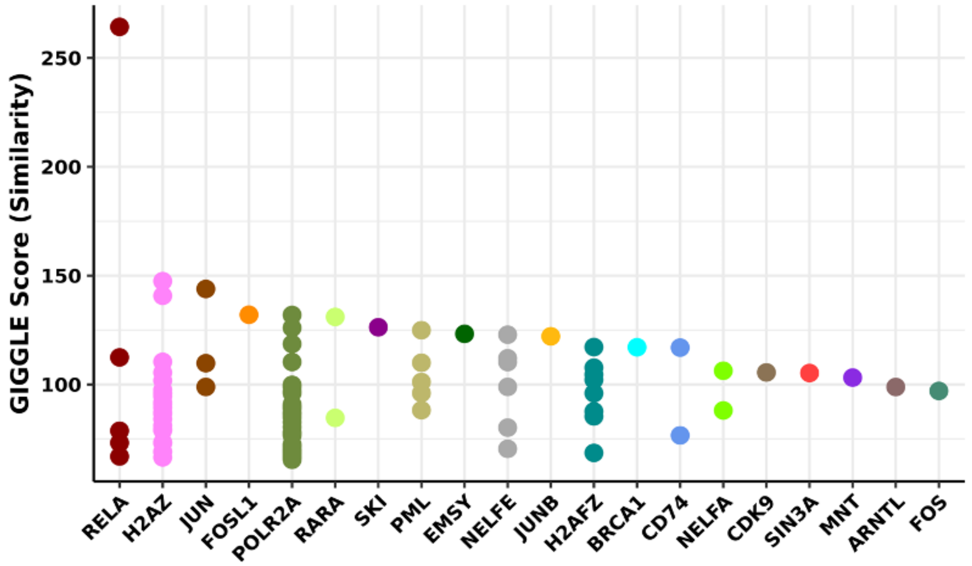


**Supplementary Figure 8. Validating the inferred TF-chromatin region linkage in MAGICAL circuits in CD14 monocytes using ChIP-seq data from the Cistrome database.** MAGICAL identified AP-1 proteins as top regulators in the circuits. During the assessment of chromatin region similarity between circuit chromatin sites and top 1000 peaks in each ChIP-seq profile (human) in the Cistrome database, JUN and FOS are top ranked too.

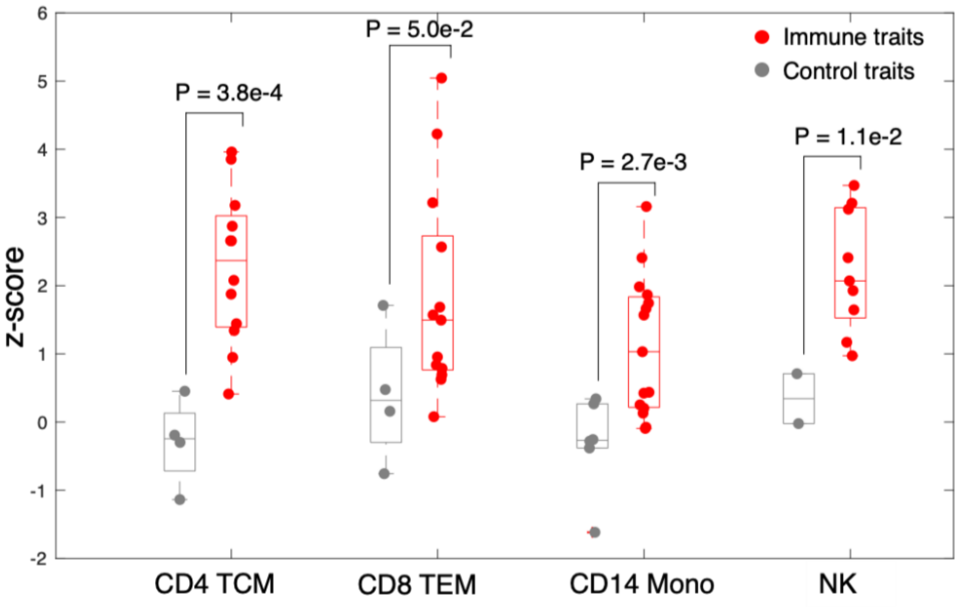


**Supplementary Figure 9. Enrichment of inflammatory disease GWAS loci in circuit chromatin sites**. Results are presented as enrichment z-score for MAGICAL-selected circuit chromatin sites in each cell type with inflammatory diseases GWAS loci (including celiac disease, crohn's disease, inflammatory bowel disease, type 1 diabetes, multiple sclerosis, primary biliary cirrhosis, rheumatoid arthritis, systemic lupus erythematosus, ulcerative colitis, psoriasis), or with GWAS loci of control diseases (Alzheimer's, ADHD, bipolar depression, Schizophrenia, Parkinson's, type 2 diabetes). Dots represent individual diseases (n = 10 for inflammatory diseases and n = 6 for control diseases). Central values represent the median z-score, the box extends from the 25th to the 75th percentile, and the whiskers extend to the maximum and minimum values no further than 1.5 times the interquartile range from the hinge. With each cell type, GWAS traits with fewer than 5 overlapped loci with circuit sites were hold out from this evaluation. The significance p-value between enrichment scores of two disease groups was assessed using two-wide Wilcoxon ranksum test.

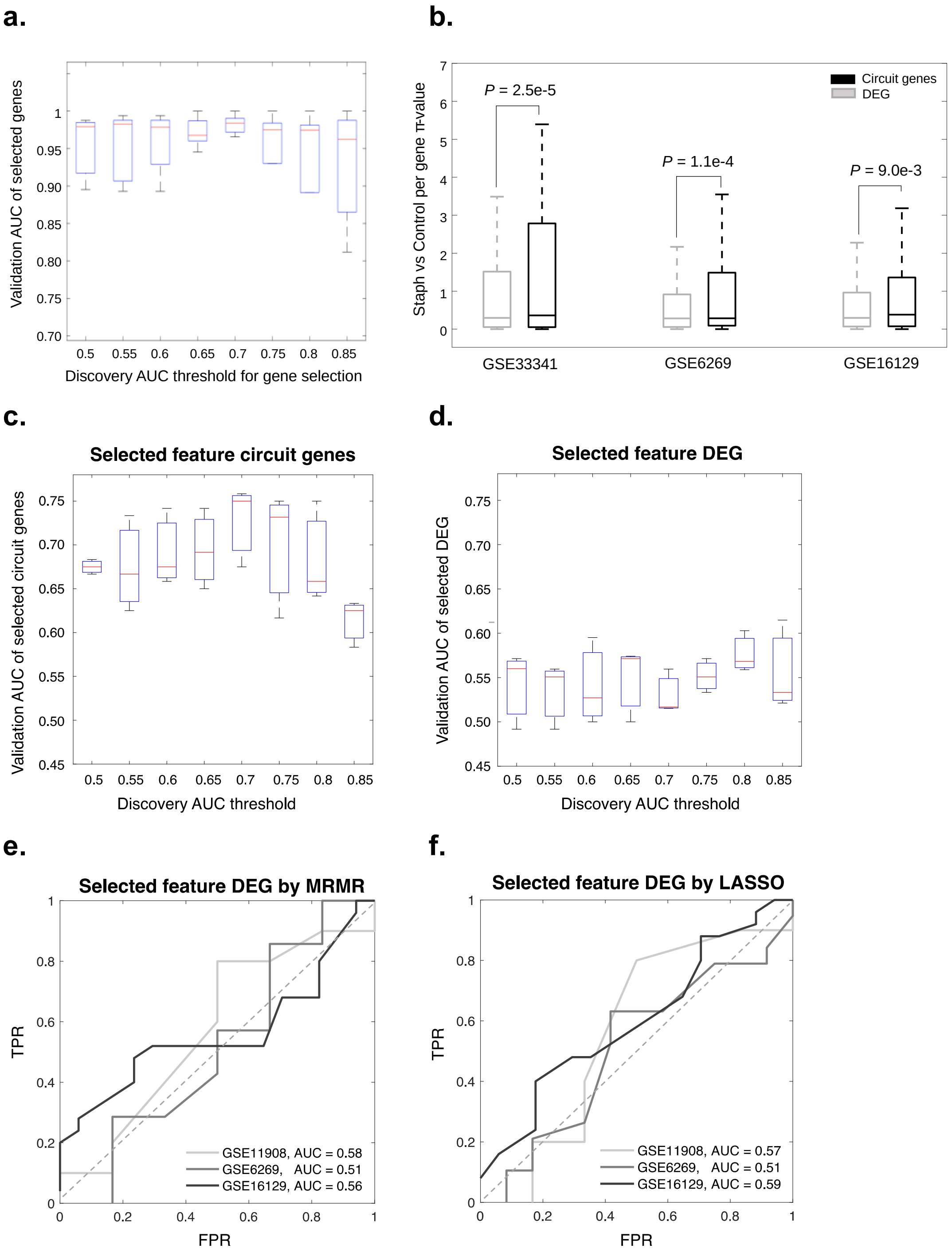


**Supplementary Figure 10. Validating circuit genes on independent microarray datasets.** (**a**) *S. aureus* vs control prediction AUCs for models that are trained with circuit genes selected above each individual cutoff (n=20 rounds of running). (**b**) *S. aureus* vs control differential expression π-values of 117 MAGICAL-identified circuit genes and 366 standard DEG in the validation microarray datasets. Significance p-value is assessed using one-side Wilconxin Ranksum test. (**c**) MRSA vs MSSA prediction AUCs for models that are trained with circuit genes selected above each individual cutoff (n=20 rounds of running). (**d**) MRSA vs MSSA prediction AUCs for models that are trained with DEG selected above the same cutoff (n=20 rounds of running). Central lines in boxplots represent the median value, the box extends from the 25th to the 75th percentile, and the whiskers extend to the maximum and minimum values no further than 1.5 times the interquartile range from the hinge. (**e**) ROC curves of predictive DEG selected by the Minimum Redundancy Maximum Relevance (MRMR) algorithm. (**f**) ROC curves of predictive DEG selected by the LASSO regression.
